# Transdiagnostic Functional Brain Alterations Across Familial and Phenotypic Risk Samples: A Meta-Analysis

**DOI:** 10.64898/2026.11.04.26367294

**Authors:** Zachary B. Millman, Debbie Burdinski, Joey Rodriguez, Tisha Chatterjea, Jennifer Urbano Blackford, Daphne J. Holt, Jacqueline A. Clauss

**Affiliations:** McLean Hospital, Belmont, MA; Harvard Medical School, Boston, MA; McGovern Institute for Brain Research, MIT, Cambridge, Massachusetts; Beth Israel Deaconess Hospital, Boston, MA; Department of Psychiatry, Massachusetts General Hospital, Boston, MA; Munroe-Meyer Institute, University of Nebraska, Omaha, NE; Maryland Psychiatric Research Center, University of Maryland School of Medicine, Catonsville, MD; Department of Psychiatry, University of Iowa Carver College of Medicine, Iowa City, IA

## Abstract

**Importance:** Psychiatric risk factors are often associated with multiple clinical outcomes. However, it remains unclear whether functional brain activation changes are shared across risk for mood, anxiety, and psychotic disorders.

**Objective:** To determine which patterns of functional brain activation are shared across risk for different psychiatric disorders, defined using phenotypic (e.g., clinical) and familial risk paradigms, relative to control samples.

**Data Sources:** PubMed and Web of Science were searched for task-based functional magnetic resonance imaging (fMRI) studies involving samples at risk for a mood, anxiety, or psychotic disorder, published between January 2000 and July 2026.

**Study Selection:** Studies published in English, with whole brain results, comparing a high-risk sample to a control sample, with a mean age of 30 years or less, were included. Of the 5,748 studies screened, 101 met selection criteria.

**Data Extraction and Synthesis:** Peak voxel coordinates from contrasts were extracted. Study quality was assessed using an adapted neuroimaging methods quality checklist. Peak coordinates were pooled within studies, and a random effects activation likelihood estimation meta-analysis was conducted using the Python package JALE.

**Main Outcomes and Measures:** Statistical maps of spatial convergence in task-related brain activation were computed across risk groups and within subgroups defined by disorder risk (mood, anxiety, or psychotic disorder risk), ascertainment paradigm (phenotypic, familial), and task domain (cognitive, affective, reward). Statistical significance was assessed using permutation-based cluster-level family-wise error (cFWE) correction.

**Results:** The studies included comprised 2534 high-risk participants and 2686 controls. Across psychosis, mood, and anxiety risk groups, there was a common area of functional activation in the left lateral prefrontal cortex (k = 319, p < .05 cFWE), reflecting greater activation in high-risk groups compared to controls. Approximately 30% of studies in each risk group contributed to this finding, reflecting both ascertainment paradigms and all three task types. Within the psychosis risk group, additional alterations were identified in the orbitofrontal cortex and anterior insula.

**Conclusions and Relevance:** Increased left lateral prefrontal activation was shared across risk groups despite methodological heterogeneity. This region supports higher-order executive functioning and undergoes prolonged maturation and this finding may represent a shared marker of vulnerability.

**Key Points:** *Question:* Are there shared differences in functional brain activation across psychiatric risk states compared with healthy controls?

*Findings:* In this meta-analysis of 101 studies involving 2534 participants at elevated risk for psychiatric disorders and 2686 control participants, participants at elevated risk had greater left lateral prefrontal cortex activation across mood, anxiety, and psychosis risk groups.

*Meaning:* Greater left lateral prefrontal activation may reflect neural processes shared across psychiatric risk states.

## Introduction

Psychiatric illness typically begins to emerge in childhood or adolescence, with risk trajectories shaped by genetic, temperamental, and clinical factors prior to onset.^1–7^ Considerable progress has been made in developing empirical “high-risk” frameworks for identifying young people at elevated risk for psychopathology, providing an opportunity to study the neural and pathogenic bases of illness.^8,9^ Critically, however, many risk factors originally identified within diagnostic silos (e.g., clinical high-risk for psychosis) are now known to confer broad vulnerability to multiple psychiatric disorders.^8^ These findings highlight the phenomenon of transdiagnostic or pluripotent risk and underscore the possibility of shared neural substrates of psychiatric vulnerability.^10,11^ Accordingly, recent years have seen an increased prominence of studies seeking to identify common risk mechanisms for psychopathology.

Historically, several complementary ascertainment paradigms have been used to operationalize risk for psychiatric disorders, including familial liability for a specific disorder (familial high-risk [FHR]), subthreshold or emerging symptoms indicating heightened clinical vulnerability (clinical high-risk [CHR]), and trait-like temperamental factors associated with later psychopathology (e.g., inhibited temperament for anxiety).^12–15^ Each approach has strengths and limitations for the study of neural substrates of risk. Familial risk paradigms offer the opportunity to study trait-like or genetically informed vulnerability in relatively asymptomatic individuals; however, their conventional focus on preadolescent samples may precede key periods of illness-relevant neurodevelopment and may overrepresent “resilient” individuals who do not progress to illness.^16–18^ In contrast, CHR approaches preferentially target adolescence and young adulthood, capturing active risk states more proximal to an evolving disease process, but commonly involve substantial symptom burden and co-occurrence of mood, anxiety, and psychosis-like phenomena.^19–23^ Although nonspecific symptom admixtures are expected during early stages of psychopathology, they complicate inferences about syndrome-specific risk mechanisms,^24,25^ particularly given the prominence of heterotypic trajectories. Critically, across ascertainment paradigms, populations defined by risk for different psychiatric outcomes are rarely examined together; thus, it remains unknown whether neural alterations attributed to illness-specific risk reflect shared underlying vulnerability.^25^

Transdiagnostic staging frameworks propose that early, nonspecific psychiatric symptoms, such as attention, anxiety, or depression symptoms, precede later more differentiated syndromes. This framework emphasizes progression along a common continuum of severity and impairment rather than categorical diagnosis.^10,26^ From this perspective, characterizing neural mechanisms associated with early stages risk syndromes may clarify both shared and divergent pathways to psychopathology and identify targets for early intervention.^27^ However, the extent to early risk syndromes share common patterns of neural alteration remains unclear.

Task-based functional MRI can be used to identify altered brain activation among subgroups of participants. In previous transdiagnostic fMRI meta-analyses of syndromal disorders, McTeague and colleagues showed that mood, anxiety, and psychotic disorders share functional impairment within multiple large-scale brain networks supporting cognitive and emotional functioning, including the prefrontal cortex (PFC), anterior insula, cingulate cortex, amygdala, hippocampus, and ventral PFC.^28–32^ Findings from individual fMRI studies of youth at risk for these disorders have been largely consistent with these findings, showing anatomically similar but more subtle patterns of aberrant functional activation relative to individuals with the disorder.^33–41^ Thus, across stages of illness, there is evidence for shared neural signatures of psychopathology, supporting the possibility of shared neural mechanisms associated with risk.

In line with this possibility are the similar trends reported between task-based fMRI studies of youth considered at risk for specific psychiatric disorders. Across cognitive and affective domains, risk-related alterations most commonly involve large-scale frontoparietal and cingulo-opercular control systems, as well as frontolimbic and frontostriatal circuitry supporting emotion, valuation, and salience processing, similar to patterns observed in established psychiatric disorders.^37,42–44^ In addition, some evidence suggests risk-specific alterations, such as cognitive-control–related impairment in psychosis risk and altered reward processing in mood disorder risk.^33,45^ Unique vulnerabilities associated with disorder-specific risk or the domain of task used to interrogate brain activation (cognitive, affective, reward-based) may therefore operate in conjunction with transdiagnostic neural mechanisms, contributing to heterogeneity within the literature.

To investigate these questions further, we conducted a meta-analysis of task-related functional MRI studies of youth selected specifically for risk of mood, anxiety, and psychotic disorders. Focusing on cognitive, reward, and affective domains, we examined differences in neural responses from control samples across multiple types of risk, including FHR, CHR, and temperament-based approaches. We hypothesized that task-related brain alterations would converge on circuits previously identified in large-scale meta-analyses of established psychopathology, including prefrontal and subcortical networks associated with cognitive, affective, and reward circuitry.

## Methods

This review was conducted using Preferred Reporting Items for Systematic Reviews and Meta-Analyses guidelines, Meta-analysis Observational Studies in Epidemiology guidelines, and neuroimaging meta-analysis best-practice recommendations (**Supplement 1**).^46,47^ The protocol was registered on the Open Science Framework (**10.17605/OSF.IO/ATNEH**) during the analysis stage (**eMethods**).

### Search and Inclusion of Studies

Articles published between January 2000 and July 2026 of task-based functional magnetic resonance imaging (fMRI) of psychiatric risk samples were reviewed. Inclusion criteria were: manuscript published in a peer-reviewed journal, task-based fMRI study, comparison of group at risk for mood, anxiety, or psychotic disorders to a healthy control group, sample mean age <30, and inclusion of whole brain results (**Figure 1**, **Supplement 2: eMethods**). Reviews, abstracts, and manuscripts not reporting primary research or not written in English were excluded. Study quality was examined by two independent raters using a 10-point checklist adapted from the Imaging Methodology Quality Assessment Checklist to the current meta-analysis sample.^43,48^ To assess interrater reliability for quality ratings, the mean absolute difference was calculated (**eMethods**).

**Figure 1:**
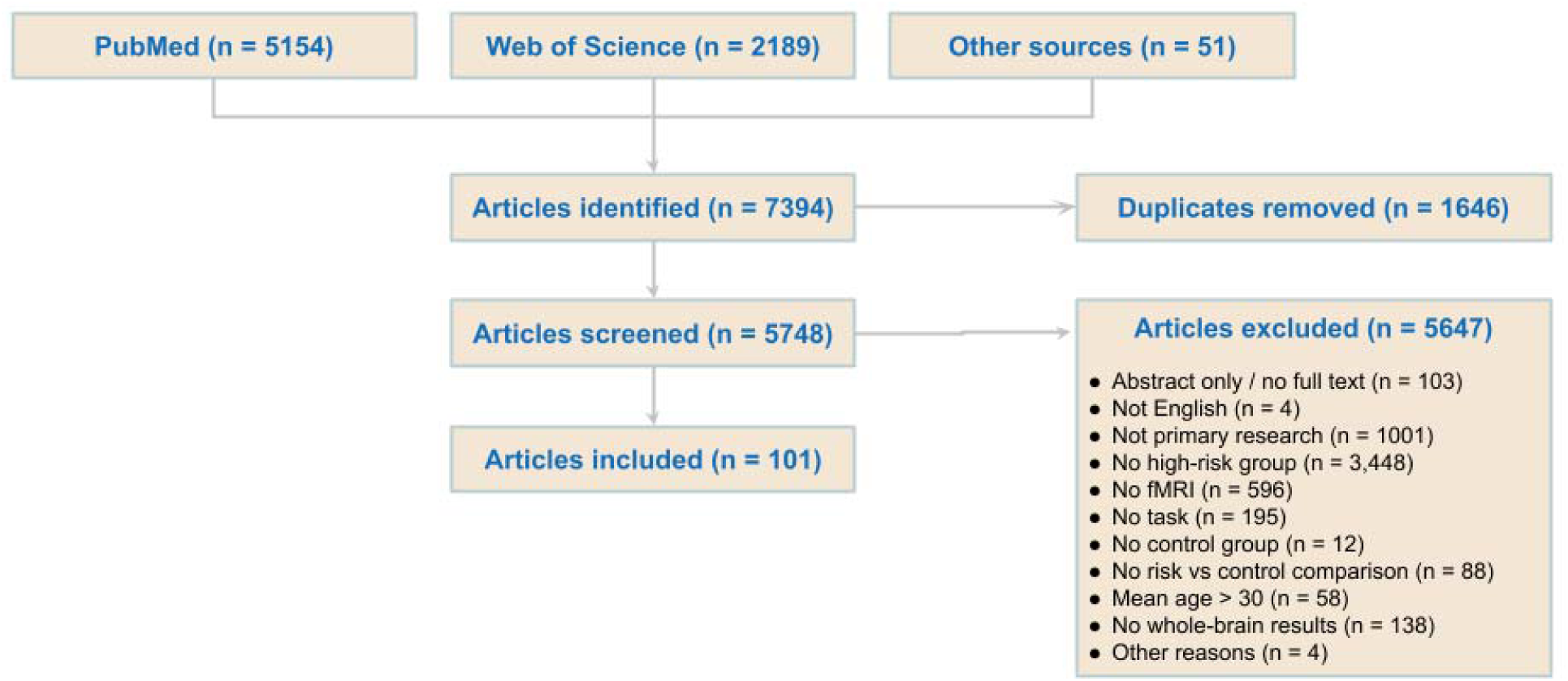
CONSORT Diagram. In total, 7394 articles were identified as being published between January 2000 and July 2026 and 5,748 were screened after removal of duplicates. 5,647 articles were excluded, most commonly for lack of a high-risk group (3,448), however other common reasons for exclusion included not primary research (1001), no fMRI (596), or no task (195). 101 articles were included in the analysis.

### Data Extraction and Preparation

Peak coordinates and study-level data were extracted by at least one reviewer and confirmed by at least one additional reviewer. Discrepancies were resolved by the first and senior author. Because our goal was to identify regions showing functional differences across groups, we extracted coordinates regardless of group-effect direction (i.e., control > risk or risk > control). This approach is consistent with prior studies and accounts for variation in contrast coding across studies (e.g., emotion > neutral vs neutral > emotion).^28,29^

Studies were grouped by the disorder for which the sample was considered at risk (“disorder-risk group”) and the empirical framework by which risk was operationalized (“ascertainment paradigm”). The latter included (1) attenuated, subclinical, and/or trait-like phenotypes (phenotypic risk; PR) and (2) a family history of the disorder (familial high-risk; FHR). The imaging tasks were categorized into cognitive, affective and reward task domains (see **eMethods)**. Peak coordinates for all contrasts reported were pooled as one study, except for one record in which data from two different risk samples were collected; these were treated here as two separate studies.^49^

### Statistical Analysis

Coordinate-based meta-analyses were performed using random effects activation likelihood estimation (ALE) implemented in the Python package JALE (https://github.com/juaml/JALE/wiki), version 0.2.7 (**eMethods**). Overall meta-analysis was conducted on all included studies to identify suprathreshold clusters of task-related activation across risk groups using a family-wise error-corrected cluster threshold 0.05 with a cluster-forming voxel-level threshold of p < .001 (cFWE). These thresholds follow recommended ALE methodology shown to provide superior control of false-positives relative to voxel-level FWE or FDR correction while retaining adequate sensitivity.^50,51^ JALE output includes a list of which studies contribute to each group-level cluster. Studies contributing to each group-level cluster were examined in post-hoc testing. To unpack group-level findings, post-hoc analyses were conducted to test for differences between studies contributing versus not contributing to a cluster. To identify smaller regions which might not have met cluster-level correction, we examined the uncorrected z-score statistical map thresholded at p < .001. Additionally, contrasts for high-risk > control and control > high risk were conducted for the overall risk meta-analysis (**eMethods, eResults**).

Exploratory meta-analyses were conducted on subsets of studies grouped by disorder risk (mood, anxiety, psychosis), ascertainment paradigm (PR, FHR), and task domain (cognitive, affective, reward; **eMethods**).

Statistical analyses were conducted in R version 4.4.1 (R Project for Statistical Computing) and Python version 3.12.6 (Python Software Foundation). Python package Nilearn and fsleyes were used for brain activation displays.^52^

## Results

### Participant and Study Characteristics

One hundred and one records were included (1.8%, 101/5748 after removal of duplicates) yielding 102 studies (one record included two different risk samples and thus was counted as two studies^49^). Studies reflecting risk for mood disorders (26 studies), psychotic disorders (65 studies), and anxiety disorders (11 studies) were classified as PR (64 studies) or FHR (39 studies) and by task domain: cognitive (52 studies), affective (33 studies), and reward-based (19 studies). See **Table 1** and **eTables 3-4**. Quality ratings averaged 9.78/10 across all studies and raters, with the absolute mean difference ratings of 0.307 (**eTable 5**).

**Table 1:**
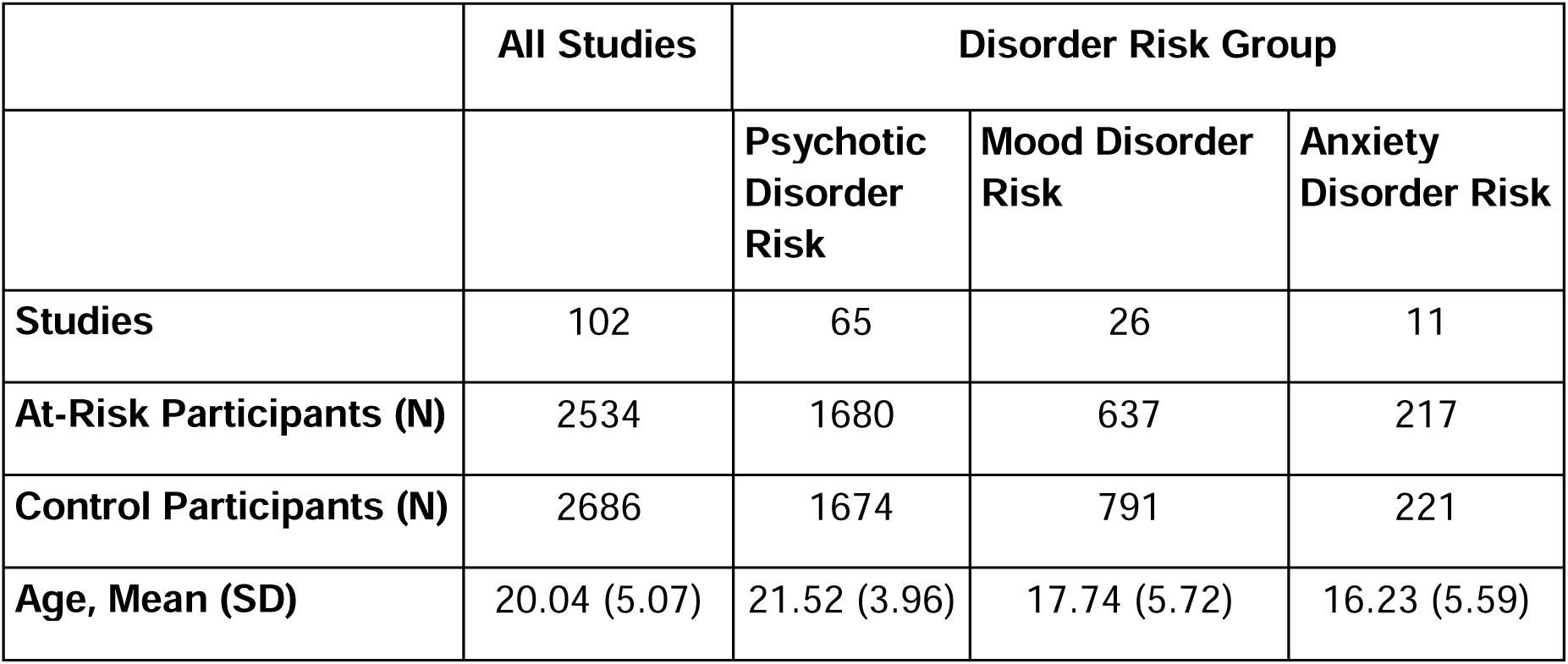
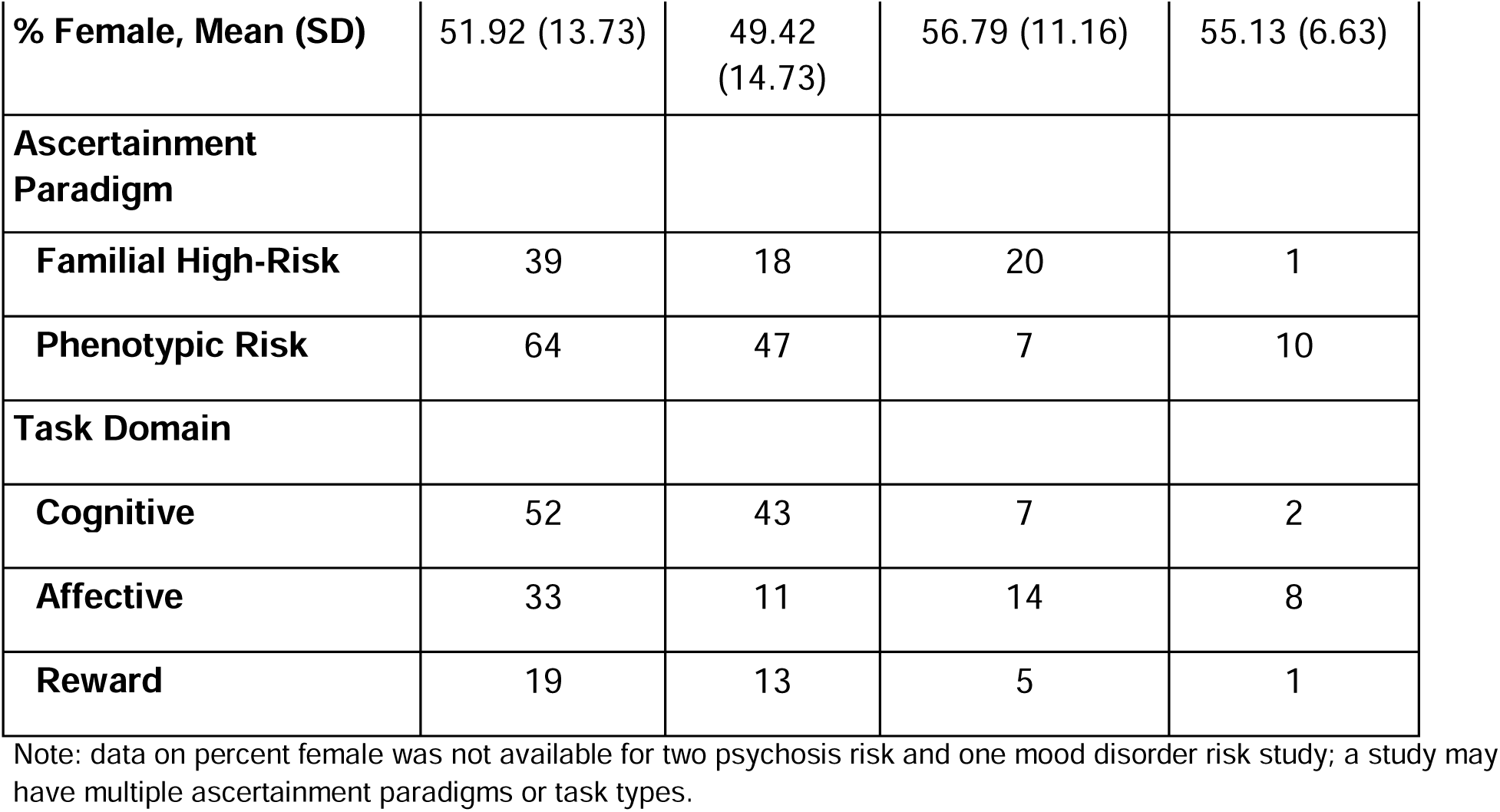
Participant Characteristics Summarized Across Disorder Risk Domains.

|  | **All Studies** | **Disorder Risk Group** |  |  |
|---|---|---|---|---|
|  |  | **Psychotic Disorder Risk** | **Mood Disorder Risk** | **Anxiety Disorder Risk** |
| **Studies** | 102 | 65 | 26 | 11 |
| **At-Risk Participants (N)** | 2534 | 1680 | 637 | 217 |
| **Control Participants (N)** | 2686 | 1674 | 791 | 221 |
| **Age, Mean (SD)** | 20.04 (5.07) | 21.52 (3.96) | 17.74 (5.72) | 16.23 (5.59) |
| **% Female, Mean (SD)** | 51.92 (13.73) | 49.42 (14.73) | 56.79 (11.16) | 55.13 (6.63) |
| **Ascertainment Paradigm** |  |  |  |  |
| **Familial High-Risk** | 39 | 18 | 20 | 1 |
| **Phenotypic Risk** | 64 | 47 | 7 | 10 |
| **Task Domain** |  |  |  |  |
| **Cognitive** | 52 | 43 | 7 | 2 |
| **Affective** | 33 | 11 | 14 | 8 |
| **Reward** | 19 | 13 | 5 | 1 |

Note: data on percent female was not available for two psychosis risk and one mood disorder risk study; a study may have multiple ascertainment paradigms or task types.

### Brain Activation Convergence across Disorder-Specific Risk Samples

*Overall Meta-Analysis (cFWE):* The overall meta-analysis across all 102 studies yielded one cluster, which was statistically significant across all risk studies. The cluster was in the left inferior and middle frontal gyrus, spanning the ventrolateral prefrontal cortex (vlPFC) and dorsolateral prefrontal cortex (dlPFC; k=319 voxels, **Figure 2A, eFigure 1**). Contributions from 30/102 studies were included and represented ∼30% of the samples in each disorder-risk group (19/65 psychosis, 7/26 mood, 4/11 anxiety; **Figure 2B, eResults**, **eFigure 2**). Post-hoc testing between the 30 studies contributing to this cluster and 72 not contributing revealed no significant study-level differences in age, percent female, disorder-risk group, ascertainment paradigm, or task domain (all p > 0.3). In a follow-up analysis, directionality of findings was examined. The overall finding of differential functional activation across studies was driven by greater activation in the risk group compared to controls (**eFigure 3**).

**Figure 2:**
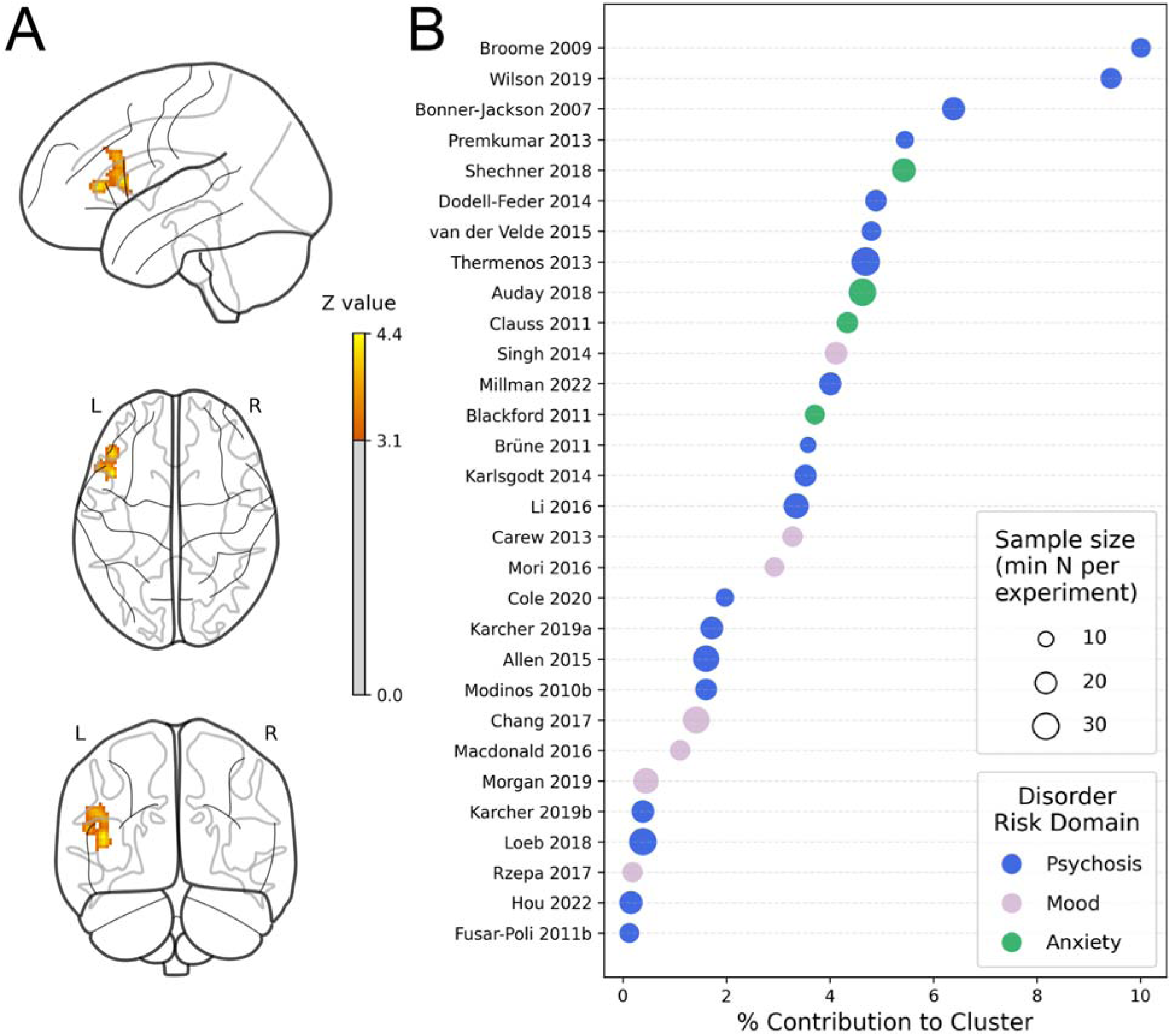
Overall Meta-Analysis: All Disorder-Risk Groups Contributed to Brain Activation Convergence (p < .05, cFWE) (A) One cluster of convergent activation across disorder-specific risk samples was identified in the left inferior and middle frontal gyrus, spanning the ventrolateral and dorsolateral prefrontal cortex (k = 319, MNI: x = -46, y = 20, z = 16). (B) 30/102 samples (disorder-risk breakdown: 19/65 psychosis; 7/26 mood; 4/11 anxiety) of comparable sample sizes contributed to the cluster.

#### Overall Meta-Analysis (non-cluster corrected)

At an uncorrected voxel-level p < .001, there were additional areas of convergence in areas of the default mode network, including the bilateral angular gyrus, left precuneus, and right parahippcampal gyrus, as well as in the bilateral frontal pole, right OFC, left lateral occipital cortex, bilateral insula, right paracingulate gyrus, and right precentral gyrus (**eFigure 4**). These results show a similar spatial distribution to the risk > control analysis (**eFigure 5).**

### Brain Activation Convergence by Disorder-Risk Group, Ascertainment Paradigm, and Task Domain

#### Psychosis Risk (cFWE)

The psychosis-risk group (65 samples) had three statistically significant clusters (**Figure 3A**, **eFigure 7**): in the right precentral gyrus, bordering the inferior and middle frontal gyrus (k = 97; 10/65 samples); right anterior insula and OFC (k = 103; 9/65 samples); and left inferior frontal gyrus (k = 175; 19/65 samples, largely overlapping with the overall meta-analysis cluster). For the two clusters which included portions of the lateral PFC, samples which contributed to these findings spanned both ascertainment paradigms (PR, FHR) and all three task domains (cognitive, reward, affective). For cluster 2, which included the right anterior insula and OFC, only cognitive and reward tasks, and only PR samples, contributed (**eFigures 8-10**).

**Figure 3:**
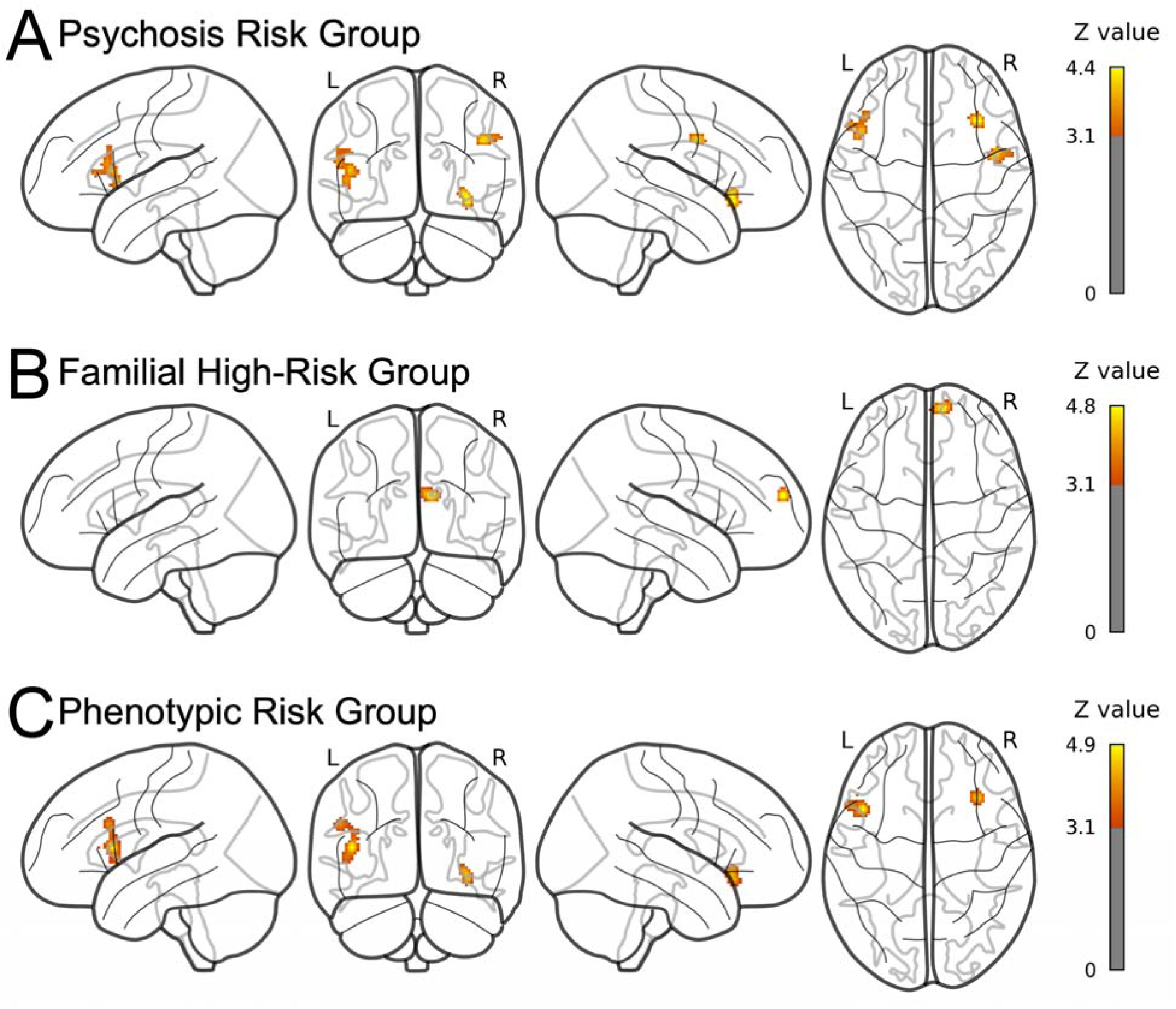
Clusters of Convergent Brain Activation Across Psychotic Disorder Risk (A), FHR (B) and PR (C) Samples (p < .05 cFWE). (A) In the psychotic disorder risk group (65 studies) three clusters of convergent activation were identified. The first cluster was in the right precentral gyrus, bordering the inferior and middle frontal gyrus and overlapping with the inferior frontal junction (k = 97 voxels; center: x = 48, y = 0, z = 34). The second cluster was in the right anterior insula and orbitofrontal cortex (k = 103 voxels; center: x = 34, y = 24, z = -6). The third cluster was in the left inferior frontal gyrus (k = 175 voxels; center: x = -46, y = 18, z = 12). (B) In the FHR group (39 studies) one cluster was identified. It was in the right frontal pole (k = 101 voxels; center: x = 10, y = 60, z = 22). (C) In the PR group (64 studies) two clusters of convergent activation were identified. The first cluster was in the right anterior insula and OFC (k = 101 voxels; center: x = 34, y = 24, z = -6). The second cluster was in the left inferior frontal gyrus (k = 203 voxels; center: x = -46, y = 18, z = 14).

#### FHR (cFWE)

The combined 39 FHR studies had one statistically significant cluster (**Figure 3B, eFigure 11**) in the right frontal pole (k = 101; 6/39 studies). Only psychosis and mood disorder risk groups, and only cognitive and affective tasks, contributed (in line with the fact that only 1/39 available studies included an anxiety risk study and 2/39 used a reward task (**eFigure 12**)). The frontal pole finding was also present in the risk > control meta-analysis (**eFigure 3**). These findings overlapped with risk > control samples (**eFigure 5, eFigure 16C**).

#### PR (cFWE)

The combined 64 PR studies had two statistically significant clusters (**Figure 3C, eFigure 13**): in the right anterior insula and OFC (k = 101; 10/64 studies); and left inferior frontal gyrus (k = 203; 20/64 studies). For cluster 1, which included the right anterior insula and OFC, all but one contributing study (anxiety risk) used a psychosis risk sample and only samples with cognitive and reward tasks contributed. For cluster 2, which included the left inferior frontal gyrus and largely overlapped with the overall meta-analysis findings, studies contributing to these findings spanned all three disorder risk groups (psychosis, mood, anxiety) and all three task types (cognitive, reward, affective; **eFigures 14-15**). These findings overlapped with psychosis risk samples and risk > control (**eFigure 16A-B).**

The remaining disorder-risk group meta-analyses (mood, anxiety) and the task type meta-analyses (cognitive, affective, reward) did not identify any statistically significant clusters after cluster-level correction. However, the meta-analyses for all disorder risk groups, both ascertainment paradigms and all task types showed convergent activation in the left dlPFC at the location of the overall meta-analysis cluster at an uncorrected voxel-level p < .001 (**Figure 4**).

**Figure 4:**
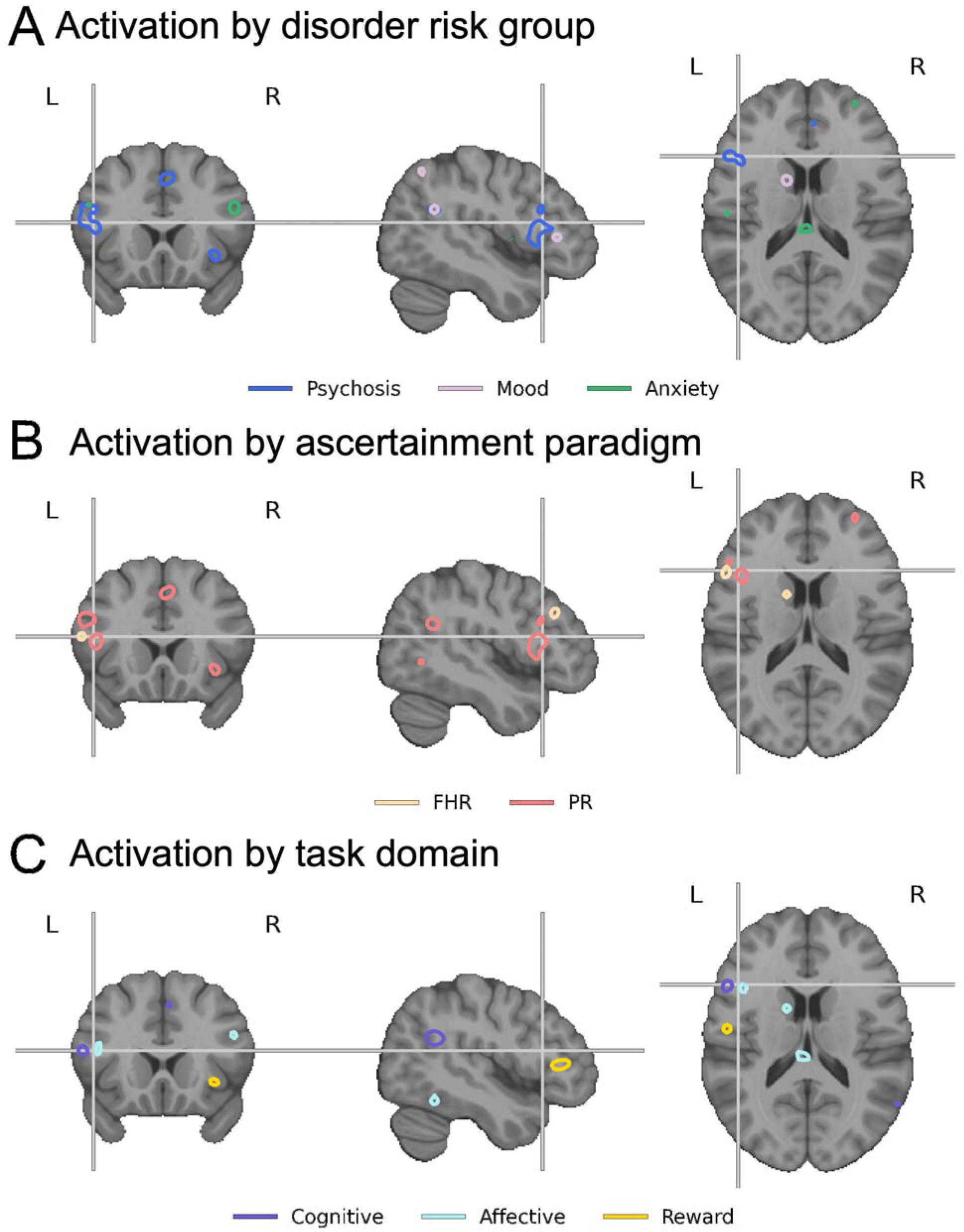
Brain Activation Convergence by Disorder-Risk Group, Ascertainment Paradigm, and Task Domain Overlap at Location of Overall Meta-Analysis Cluster. Contours of brain activation convergence at voxel wise p < .001 uncorrected in individual (A) disorder risk domain, (B) risk type, and (C) task type meta-analyses at the location of the overall meta-analysis cluster (center indicated by cross).

#### Psychosis Risk (non-cluster corrected)

The psychosis-risk meta-analysis revealed additional convergent activation in regions of the default mode network (left angular gyrus and left precuneus), as well as portions of the frontal cortex involved in higher order processing (left superior frontal gyrus, right paracingulate gyrus, and right frontal pole; **Figure 4A; eFigure 17**).

#### Mood Disorders Risk (non-cluster corrected)

The mood-risk meta-analysis revealed convergent activation in portions of the visual cortex (bilateral occipital cortex and left occipital pole), portions of the default mode network (left lateral parietal cortex and angular gyrus), as well as in somatosensory regions (postcentral gyrus), parts of the striatum (the left caudate), and the right frontal pole (**Figure 4A; eFigure 17**).

#### Anxiety Disorders Risk (non-cluster corrected)

The anxiety-risk meta-analysis revealed convergent activation in regions involved in social-affective processing (left middle and superior temporal gyrus, left insula, left frontal pole, and right inferior and middle frontal gyrus), as well as left the cerebellar hemisphere and left caudate tail (**Figure 4A**; **eFigure 17**).

#### FHR (non-cluster corrected)

The FHR meta-analysis revealed convergent activation in default mode network regions (left inferior frontal gyrus, left anterior cingulate gyrus, right frontal pole and right angular gyrus), frontoparietal network regions (left middle frontal gyrus), visual/dorsal attention network regions (left and right lateral occipital cortex) and somatomotor network regions (right planum polare; **Figure 4B; eFigure 18**).

#### Phenotypic Risk (non-cluster corrected)

The PR meta-analysis revealed convergent activation in default mode network regions (left inferior and superior frontal gyrus, left middle temporal gyrus, and left and right angular gyrus), frontoparietal network regions (left middle frontal gyrus, right frontal pole, right orbitofrontal cortex and right paracingulate gyrus), ventral attention network (left precentral gyrus, left and right insula), dorsal attention network (right precentral gyrus) and visual network (right cuneus; **Figure 4B; eFigure 18**).

#### Cognitive Task (non-cluster corrected)

The cognitive task meta-analysis showed convergent activation in prefrontal regions (left inferior frontal gyrus and right OFC), the parietal cortex (bilateral angular gyrus, right superior parietal lobule, and right precentral gyrus) as well as the right paracingulate gyrus (**Figure 4C; eFigure 19**).

#### Affective Task (non-cluster corrected)

The affective task meta-analysis showed convergent activation in prefrontal regions (right pars opercularis, left inferior and middle frontal gyrus, and bilateral frontal pole), left middle temporal gyrus, visual cortex regions (left fusiform gyrus, right precuneus, right lateral occipital cortex), motor regions (left precentral gyrus), left caudate, and limbic regions (right cingulate and paracingulate gyri, left OFC, and right parahippocampal gyrus; **Figure 4C; eFigure 19**).

#### Reward Task (non-cluster corrected)

The reward task meta-analysis showed convergent activation in prefrontal regions (left pars opercularis, left inferior and superior frontal gyri and right frontal pole), limbic regions (left parahippocampal gyrus, and right paracingulate gyrus), right insula, left lateral occipital cortex, right supramarginal gyrus, and subcortical regions (bilateral thalamus and right caudate; **Figure 4C; eFigure 19**).

## Discussion

Identifying transdiagnostic neural substrates of risk has become a central focus of psychiatric research, but progress has been constrained by the field’s tendency to study risk groups in isolation and to rely on narrative comparisons across studies. The present meta-analysis aimed to address this gap, revealing convergent prefrontal functional alterations across diverse disorder-risk groups and ascertainment paradigms, with both shared and selective patterns that have implications for developmental neuroscience, transdiagnostic models, and research design.

The overall pattern suggesting altered vlPFC, dlPFC, and salience network activation is consistent with meta-analyses of established psychopathology showing disruption within a “multiple demand” network supporting cognitive control^7,28,29,53^. McTeague and colleagues identified common dysfunction across psychotic and nonpsychotic disorders in regions including the vlPFC, dlPFC, anterior insula, and paracingulate cortex during cognitive control^29^. Our results suggest altered functional activation of these regions may reflect transdiagnostic psychiatric vulnerability rather than a marker or consequence of established illness.

In contrast, a psychosis-risk cluster encompassing the anterior insula and orbitofrontal cortex showed selective contributions from phenotypic risk samples and from cognitive and reward paradigms. Abnormalities within this network, which supports salience detection and value-based decision-making^54^, have been consistently demonstrated in established psychosis^28,55^. It is possible that more selective alterations associated with psychosis risk emerge as a “second wave” of vulnerability, superimposed on preexisting transdiagnostic risk,^56^ during the emergence of more complex or psychosis-related psychopathology.

Analyses restricted to mood- and anxiety-risk studies revealed no convergent clusters at cluster corrected thresholds, potentially reflecting limited power. At a more liberal voxel-level threshold, however, both groups converged in the left dlPFC, overlapping the primary transdiagnostic cluster. Prior meta-analyses of established mood and anxiety disorders report robust abnormalities in prefrontal, parietal, and striatal regions^29,53,57^. These findings suggest that prefrontal control alterations may extend to risk groups but with smaller effects or greater heterogeneity that limit detection under more stringent thresholds. Uncorrected analyses nonetheless revealed more distributed involvement –– visual, default mode, and striatal regions in mood-risk studies and social-affective, insular, and frontotemporal regions in anxiety-risk studies –– suggesting more diffuse alterations than the robust prefrontal-salience convergence seen in psychosis-risk samples.

The common functional alterations within the vlPFC and dlPFC have implications for developmental neuroscience. These regions support higher-order cognitive abilities broadly implicated in psychopathology (e.g., working memory, language, decision-making^58,59^) and exert top-down control over subcortical affective and reward circuitry^60,61^. The PFC is among the last brain regions to mature during development^62^, implicating a protracted developmental window of vulnerability to environmental insults and divergent neuromaturational trajectories^63,64^. The dlPFC is also the primary target for repetitive TMS in depression and related conditions^65^, raising questions about whether preexisting vulnerability shapes treatment response and offering a rationale for PFC-targeted cognitive training in high-risk populations^66^.

The present findings support transdiagnostic staging frameworks, which propose that early psychopathology reflects shared neurobiological vulnerability that later differentiates into syndrome-specific patterns^10^. They also align with dimensional paradigms including the Research Domain Criteria and the Hierarchical Taxonomy of Psychopathology^67,68^, which treat neural engagement as continuous variation along neurobehavioral and clinical dimensions rather than categorical pathology. Integrating these frameworks, though challenging, may help delineate where shared vulnerability progresses to syndrome-specific circuitry.^69^

A significant implication of this study is that neuroimaging studies focusing on single-disorder risk groups may conflate disorder-specific vulnerability mechanisms with more general features of psychopathology or risk. This problem is well-recognized in the established illness literature^70^ but is underappreciated in risk research^25^, despite consensus that illness presentations and mechanisms have less specificity before onset than afterward^26,71–73^. The present findings extend clinical^22,74^, neuroanatomical^75^, and genetic evidence^76^ of shared profiles across risk groups to the domain of functional brain activation. Although pathways to common localization may differ and direct risk-group comparisons were precluded by data availability, the results provide an indirect test of specificity and underscore the need for transdiagnostic risk paradigms.

Several limitations warrant consideration. First, the number of anxiety-risk studies was relatively small. This group nevertheless contributed proportionally to overall convergence, suggesting the main findings were not driven solely by mood and psychosis risk. Second, uneven distribution of task types across disorder-risk classes limited the ability to isolate effects of these characteristics, highlighting the need for more balanced designs in future work. Third, medication status varied across studies; although treatment rates were generally lower than in studies of established disorders^28,77^, its influence on these results cannot be ruled out. More broadly, coordinate-based meta-analysis prioritizes spatial convergence of reported peaks, potentially underrepresenting smaller or more heterogeneous effects and precluding effect size estimation and unreported null findings^78^. Prospective designs are needed to address these constraints.

In conclusion, this meta-analysis identified convergent functional alterations in the dlPFC and vlPFC across disorder-risk groups, ascertainment paradigms, and task domains, with additional salience and orbitofrontal alterations in symptom-based psychosis-risk cohorts and cognitive and reward tasks. The findings implicate a transdiagnostic neural substrate of psychopathology risk while providing evidence for specificity in some contexts, a pattern consistent with contemporary transdiagnostic and developmental accounts of psychopathology.

## Data Availability

Source code and data will be made available publicly upon publication; see Supplementary Material. Source code and data will be made available to reviewers and editors during the review process as requested.

## Author Contributions

Conceptualization: ZBM, JUB, DJH, JAC

Methodology: ZBM, DB, JAC

Software: DB, JAC

Investigation: ZBM, DB, TC, JR, JAC

Formal Analysis: DB, JAC

Data Curation: ZBM, DB, TC, JR, JAC

Writing – original draft: ZBM, DB, JAC

Writing – final draft: ZBM, DB, TC, JR, JUB, DJH, JAC

Supervision: JUB, DJH, JAC

## Additional Contributions

The authors thank Dylan Cugley and Katherine Dokholyan for their early contributions to data extraction and quality assessment, Veronika Muller for her guidance on use of ALE analysis and software, and Maya Schumer for her recommendations on analytic strategies and introduction to the JALE investigators.

