## Supplement 2 for "Transdiagnostic Functional Brain Alterations Across Familial and Phenotypic Risk Samples: A Meta-Analysis"

**Protocol Registration:** [**10.17605/OSF.IO/ATNEH**](https://doi.org/10.17605/OSF.IO/ATNEH)

**Contents**

**eMethods**: Search and Inclusion of Studies, Inclusion and Exclusion Criteria, Quality Assessment, Data Extraction, Data Preparation, Peak Coordinates, Statistical Analysis

**eResults**: Convergence in Brain Activation across Disorder-Risk Groups; Brain Activation and Convergence by Disorder-Risk Group, Ascertainment Paradigm, and Task Domain; Sensitivity Analyses

**eTable 1**: Search Terms by Database

**eTable 2**. Imaging Methodology Quality Assessment Checklist Adapted for Psychiatric Risk Samples

**eTable 3**: Participant Characteristics by Study

**eTable 4**: Task Characteristics by Study

**eTable 5**: Quality Assessment

**eFigure 1**: Brain Activation Convergence (cFWE corrected) in the Overall Meta-Analysis across All Samples.

**eFigure 2**: All (A) Task Domains and (B) Ascertainment Paradigms Contributed to Brain Activation Convergence in the Overall Meta-Analysis across All Samples

**eFigure 3:** Overall Meta-Analysis across All Samples Driven by Risk > Control (cFWE corrected).

**eFigure 4**: Brain Activation Convergence (not cFWE corrected) in the Overall Meta-Analysis across All Samples

**eFigure 5:** Risk > Control Brain Activation Convergence (not cFWE corrected) in the Overall Meta-Analysis across All Samples

**eFigure 6:** Control > Risk Brain Activation Convergence (not cFWE corrected) in the Overall Meta-Analysis across All Samples

**eFigure 7**: Brain Activation Convergence (cFWE corrected) in the Psychotic Disorder Risk Meta-Analysis

**eFigure 8**: All (A) Task Domains and (B) Ascertainment Paradigms Contributed to Brain Activation Convergence in Cluster 1 in the Psychotic Disorder Risk Meta-Analysis

**eFigure 9**: (A) Cognitive and Reward Tasks and (B) Symptom and Trait-Based Risk Samples Contributed to Brain Activation Convergence in Cluster 2 in the Psychotic Disorder Risk Meta-Analysis

**eFigure 10**: All (A) Task Domains and (B) Ascertainment Paradigms Contributed to Brain Activation Convergence in Cluster 3 in the Psychotic Disorder Risk Meta-Analysis

**eFigure 11**: Brain Activation Convergence (cFWE corrected) in the FHR Meta-Analysis

**eFigure 12**: (A) Psychosis and Mood Risk Samples and (B) Cognitive and Affective Tasks Contributed to Brain Activation Convergence in the FHR Meta-Analysis

**eFigure 13**: Brain Activation Convergence (cFWE corrected) in the PR Meta-Analysis

**eFigure 14**: (A) Psychosis and Anxiety Risk Samples and (B) Cognitive and Reward Task Domains Contributed to Brain Activation Convergence in Cluster 1 in the PR Meta-Analysis

**eFigure 15**: All (A) Disorder-Risk Group Samples and (B) Task Domains Contributed to Brain Activation Convergence in Cluster 2 in the PR Meta-Analysis

**eFigure 16**: Substantial Overlap of Between (A) the Overall Meta-Analysis with All Samples Cluster, the Psychosis-Risk Meta-Analysis Cluster 3 and the PR Meta-Analysis Cluster 2, as well as between (B) the Psychosis-Risk Meta-Analysis Cluster 2 and the PR Meta-Analysis Cluster 1, and (C) Overlap Between the Familial High-Risk Meta-Analysis, the Overall Risk > Control Analysis, and the Overall Meta-Analysis Across All Samples.

**eFigure 17**: Brain Activation Convergence (not cFWE corrected) in the Three Disorder-Specific Risk Meta-Analyses.

**eFigure 18**: Brain Activation Convergence (not cFWE corrected) in the Two Disorder-Risk Group Meta-Analyses

**eFigure 19**: Brain Activation Convergence (not cFWE corrected) in the Three Task Domain Meta-Analyses

**eReferences**

**eMethods**

Protocol Registration

During the search stage of this review, we prepared the written protocol for registration with PROSPERO, with preregistration attempted prior to completion of screening. The submission did not result in a confirmed registration, however, and this incompleteness was identified after data analysis had been begun. To clarify why the original submission had not been registered, a second attempt was made, which also did not result in registration confirmation. We therefore opted to register the review with the Open Science Framework (OSF).

Given that the OSF registration was completed after major analyses were complete, the registration is considered largely retrospective, documenting original study aims and methodological choices as well as procedures implemented during the project. Analyses planned after inspecting the initial findings are identified as post hoc in the registration and in the manuscript.

Search and Inclusion of Studies

PubMed and Web of Science were searched for articles published between January 2000 and July 2026 that involved task-based functional neuroimaging (fMRI) of psychiatric risk samples. References from included studies, review articles and meta-analyses were examined to identify additional studies for inclusion. Study inclusion criteria included: task-based fMRI, inclusion of a high-risk group for psychosis, anxiety disorders, or mood disorders; comparison of at-risk sample versus healthy control.

Specifically, studies underwent screening if they utilized task-based fMRI and indicated the sample being studied was at-risk for anxiety, mood or psychotic disorders (see Inclusion and Exclusion Criteria below). Studies were screened by at least one rater (ZBM or JAC). Included studies were reviewed by both ZBM and JAC and conflicts were decided by consensus. To assess interrater reliability, the first 100 records from the second half of the search results (alphabetized by first author surname) were assessed by both raters; agreement was 100%. Search terms are presented in eTable 1.

**eTable 1: Search Terms by Database**

| **Database** | **Search Terms** |
| --- | --- |
| Web of Science | (“functional MRI” OR fMRI) AND (depression OR bipolar OR psychosis OR anxiety) AND risk  (“functional MRI” OR fMRI) AND (‘‘inhibited temperament’’ or “behavioral inhibition”)  ("functional MRI" OR fMRI) AND ("psychotic experiences" OR "psychotic symptoms" OR "psychotic-like experiences" OR "psychoticlike symptoms" OR "nonclinical psycho*" OR "subclinical psycho*" OR "psychosis spectrum") |
| PubMed | (“functional MRI” OR fMRI) AND (depression OR bipolar OR psychosis OR anxiety) AND risk  (“functional MRI” OR fMRI) AND (‘‘inhibited temperament’’ or “behavioral inhibition”)  ("functional MRI" OR fMRI) AND ("psychotic experiences" OR "psychotic symptoms" OR "psychotic-like experiences" OR "psychoticlike symptoms" OR "nonclinical psycho*" OR "subclinical psycho*" OR "psychosis spectrum") |

Inclusion and Exclusion Criteria

1. Full text available
2. Published in English
3. Empirical study using task-based functional MRI were included.
   1. Review articles and meta-analyses were excluded.
   2. Other imaging modalities were excluded, including positron emission tomography (PET), single photon emission tomography (SPECT), and magnetoencephalography (MEG).
   3. Resting state functional MRI studies were excluded.
4. High-risk groups: Procedures must have involved identifying a group at elevated risk for developing a mood, anxiety, or psychotic disorder. Operationalization was based on the prior literature; the following were included:
   1. Familial risk (i.e., first- or second-degree relative with the diagnosis of interest for that study);
   2. Subclinical symptoms or syndrome (e.g., subclinical depressive or manic symptoms in the absence of lifetime mood disorder; clinical high-risk for psychosis or elevated psychotic experiences);
   3. Trait-like presentations (i.e., elevated schizotypy, inhibited temperament or behavioral inhibition in anxiety disorders literature).
5. High-risk groups: The following operationalizations of risk were excluded:
   1. Risk defined solely through a dimensional measurement without case-control comparison (e.g., a study of neural engagement associated continuously with subclinical depressive symptoms).
   2. Risk defined by an individual genetic polymorphism (i.e., 22q11).
   3. Risk defined solely by environmental factors such as trauma exposure.
   4. Animal models of psychopathology.
6. Comparison made between a group at high risk with a control group at lower risk.
7. Reporting of peak voxels for differences between the high-risk and control groups reported in a standardized space (MNI or Talairach).
   1. Only whole brain analyses were included.
   2. Studies which only reported region-of-interest results were excluded.
8. Average age of subjects 30 years or lower.

Quality Assessment

To assess the quality of scientific reporting and methodology among the included studies, quality, following prior work the Imaging Methodology Quality Assessment Checklist (IMQAC) was adapted to fit psychiatric risk samples.^1,2^ The IMQAC evaluates studies based on 10 categories, each scored 0 (criteria unfulfilled), 0.5 (criteria partially fulfilled), or 1 (criteria fulfilled), for a total possible score of 10. The instrument was adapted by adding contextualized definitions and by operationalizing the IMQAC criteria. The adapted instrument is presented in eTable 2; non-boldfaced text (itemized criteria and definitions) within each category represents the present adaptation

**eTable 2. Imaging Methodology Quality Assessment Checklist Adapted for Psychiatric Risk Samples**

**IMAGING METHODOLOGY QUALITY ASSESSMENT CHECKLIST**

**Score:** (0/0.5/1)

**Category 1: Subjects**

1. **High-risk participants were evaluated prospectively, specific diagnostic criteria were applied, relevant psychiatric and medical illnesses were excluded, and demographic data were reported.**

- Participants were assessed prior to having ever developed the disorder of interest.
- Operationalized high-risk criteria were applied using validated instruments.
  - For familial high-risk studies, diagnosis of the affected relative (e.g., parent) was determined using a validated instrument via direct interview with the relative (e.g., MINI, SCID) or indirect interview with the youth participant (e.g., FIGS).
- Exclusion of “relevant psychiatric illnesses” is restricted to substance use disorders (e.g., studies are not penalized for allowing co-occurring disorders, except substance use disorders).
- Exclusion of medical illnesses is restricted to neurological disorders (e.g., traumatic brain injury, epilepsy).
- Age and sex of participants reported.

2. **Healthy comparison subjects were evaluated prospectively, psychiatric and medical illnesses were excluded, and demographic data were reported**

- Healthy control participants did not have the diagnosis of interest within the high-risk sample.
- Medical exclusions and demographic reporting same as item #1.

3. **Important variables (e.g., age, gender, intelligence quotient, i.e., IQ, handedness, motion, quality control) were checked, either by stratification or statistically.**

- All reported variables listed above were shown separately by group.

4. **Sample size per group > 10**.

**Category 2: Methods for image acquisition and analysis**

5. **All neuroanatomic and functional measurements were taken without considering group assignment or participant identity.**

- The same neuroimaging and data processing protocol was used across study groups and demographic factors.

6. **Magnet strength > 1T.**
7. **The imaging technique used was clearly described so that it could be reproduced. TR, TE, #coils, clearly described paradigm (length, # of stimuli, randomization of stimuli).**

- TR, TE, voxel size, smoothing kernel, stimulus duration, and acquisition time are reported.
- Randomization/pseudo-randomization of stimuli is described in situations in which there is reason to believe the ordering of stimuli would confound the main outcome parameters.

8. **Measurements of task parameters were clearly described so that they could be reproduced.**

- For event-related tasks this may refer to the construction of regressors based on time stamps of relevant task events.
- For block design tasks this may refer to the fitting of regressors to BOLD time series data for each block.

**Category 3: Results and conclusions**

9. **Statistical parameters for significant, and important non-significant, differences were provided.**

- Test statistics and/or measures of effect size are reported for the primary analysis.
- Important non-significant differences refer to the primary analysis.

10. **Conclusions were consistent with the results obtained and the limitations were discussed.**

- Study limitations are discussed and conclusions are reasonable.

**TOTAL:** /10

Data Extraction

Participant characteristics, study characteristics, and peak voxel coordinates from whole brain analyses of interest (directional group contrasts: at-risk > control and control > at-risk; non-directional omnibus tests involving group: main effects and interactions) were extracted. All data extraction was verified by at least one additional team member. Any conflicts were resolved through review by first or senior authors. Coordinates from region of interest analysis or with small volume correction were excluded so as to reduce bias based on *a priori* lower statistical thresholds.

Data Preparation

*Sample Classification*

Samples were first grouped by the family of psychiatric diagnoses defining the risk sample (i.e., mood, anxiety, or psychotic disorder risk; “disorder-risk group”). Studies investigating risk for unipolar or bipolar mood disorders were combined given the limited number of studies available support independent analyses.

Samples were further organized broadly according to how risk for a given disorder was operationalized (i.e., “ascertainment paradigm”; **eTable 3**). The present study was intentionally designed to test the hypothesis that distinct traditions across psychopathology literatures may converge on a similar pattern of findings. Thus, we anticipated heterogeneous operationalizations of risk both within fields (e.g., within studies of psychosis risk) and between them (e.g., between studies of psychosis risk and mood disorder risk). The goal of our classification scheme was to provide a common analytic framework for this heterogeneous literature while preserving meaningful distinctions between broad risk constructs.

With this in mind, we organized samples according to whether risk was determined based on familial liability (FHR) or symptom or trait-based risk (phenotypic risk; PR). FHR samples were defined by a positive family history (first- or second-degree relative) of the disorder in question and did not require the expression of any symptom, behavior, or trait. PR samples were defined by observable signs or reported mental phenomena presumed within their respective fields to reflect the phenotypic expression of an underlying vulnerability to the disorder in question. For study- or field-level criteria incorporating both familial and phenotypic factors, we classified samples as PR when the expression of the relevant phenotype was required for positive risk status (i.e., when familial liability alone was insufficient for determining risk).

Psychosis-risk samples classified as phenotypic risk included clinical high-risk, elevated psychotic experiences, and elevated levels of schizotypy. Mood-risk samples classified as phenotypic risk included subthreshold depressive symptoms, subthreshold manic syndromes, and for one study,^3^ depressive or attention-deficit/hyperactivity syndromes plus a biological parent with bipolar disorder. Anxiety-risk samples classified as phenotypic risk included elevated behavioral inhibition or inhibited temperament.

These higher-order classifications were not intended to imply orthogonality between the resulting groups or equivalence with respect to factors such as age, disorder transition rates, sampling methodology, assessment methods, or co-occurring clinical features. Rather, heterogeneity in the definition and ascertainment of risk was itself a primary motivation for the present review. We note that although the field of transdiagnostic risk phenotyping is emerging, no standards currently exist that are sufficiently established and validated for application across the literatures reviewed here. The classification scheme developed for the present study therefore provided an analytic framework for assessing convergence and divergence across risk literatures focusing on distinct end-point diagnoses and unique ascertainment paradigms.

*Task Domain Classification*

The neuroimaging tasks were categorized into cognitive, affective and reward domains (**eTable 3**). When a task design included features potentially relevant to more than one domain, the task was classified according to the primary construct the task was designed to assess. For example, a task designed to elicit negative affect using forced-fail mental arithmetic and evaluative feedback was classified into the affective domain; a task designed to elicit reward processes through probabilistic reinforcement learning was classified into the reward domain.

Peak Coordinates

All peak coordinates from eligible contrasts within a given analytic sample were pooled into a single study. Choi et al. included data from two separate risk samples and therefore treated as two separate studies.^4^

Statistical Analysis

Briefly, activation likelihood estimation (ALE) estimates spatial convergence of brain activation across neuroimaging contrasts for each study. Each study contributes peak activation voxel coordinates.^5–10^ Within each study, peaks are then modeled using a 3-dimensional Gaussian kernel to reflect spatial uncertainty. The kernel is weighted by sample size, with smaller sample sizes resulting in a wider kernel with less intensity, reflecting increased spatial uncertainty, and larger sample sizes resulting in a narrower kernel with greater intensity, reflecting less spatial uncertainty.^5^ Gaussians are combined using a voxel-wise maximum to yield a modeled activation map for each study. Modeled activation maps are then combined across studies, yielding a group-level ALE statistic map, in which the voxel-wise ALE value reflects both the number and the spatial specificity of contributing studies. Assuming spatial independence across studies, a null distribution is then analytically derived via non-linear histogram integration, allowing for efficient random-effects inference on spatial convergence.^6^ Results are corrected for multiple comparisons at the cluster-level using permutation-based cluster-level family-wise error (cFWE) correction. First clusters are identified using a voxel-wise cluster-forming threshold. Then each cluster’s cFWE-corrected p-value is derived by comparing the observed cluster size to a null distribution of cluster sizes from repeated ALE analyses on the same studies but with spatially randomized foci.^6^

Following best practices, we used the smaller sample size across at-risk and control samples for Gaussian kernel weighting, set the cluster-forming threshold to p < .001, the number of simulations to create a null distribution of cluster sizes to 5000, and the significance threshold after cFWE correction to p < .05. For the overall analysis, we did not stratify by the original modeling set-up (at-risk > control; control > at-risk; non-directional tests involving group), as our aim was to identify areas of altered functional activation, consistent with prior meta-analyses.^11,12^

*Directionality-Stratified Analyses:* Contrasts were separated by directionality (i.e., risk > control vs control > risk). Task contrasts were harmonized so that the direction of effect consistently reflected greater cognitive load for cognitive tasks, greater reward value/less loss for reward tasks, or greater emotional salience/affective intensity for affective tasks; contrasts were reversed when necessary.

*Post Hoc Testing:* Our overall meta-analysis was conducted on all included studies to identify suprathreshold clusters of task-related activation across psychiatric at-risk groups. To unpack group-level cluster findings, we examined which studies contributed to a cluster, including sample sizes and relative contributions to the cluster’s summed ALE values. Post-hoc analyses then tested for systematic differences between studies contributing to a cluster and those not contributing. Wilcoxon rank-sum tests compared mean age and percent female. Fisher’s exact tests compared risk status definition, disorder risk domain and task type. Data on sex was not available for three studies. Finally, the uncorrected z-score statistical map thresholded at p < .001 was inspected to identify smaller areas of functional difference. Cluster-based thresholds restrict to only larger clusters and may bias results towards larger areas and away from subcortical areas or smaller functional regions.

*Exploratory Analyses:* Eight exploratory meta-analyses were conducted on subsets of studies grouped by disorder domain (mood, anxiety, psychosis), by risk type (FHR, PR) or by task type (cognitive, affective, reward). Suprathreshold clusters were examined in terms of contributing studies’ sample size and ALE contribution, and uncorrected z-score statistical maps thresholded at p < .001 were inspected.

*Sensitivity Analyses:* Two sensitivity analyses were conducted for the overall meta-analysis. (1) To assess the impact of sample size specifications on modeling spatial uncertainty, the combined sample size of risk and control groups was used to derive Gaussian kernel width. (2) To consider shared variance of overlapping samples, all studies were examined for any statements of overlap between samples. If a study included an explicit statement regarding overlap of the risk sample with another study, the peak coordinates were merged into one study, with the sample size taken to be that of the smallest at-risk or control group in the studies.

Standard heterogeneity statistics used in effect-size–oriented meta-analyses are not applicable to coordinate-based meta-analysis including ALE, given that these methodologies rely on peak coordinates rather than effect size estimates and between-study variance statistics. Thus, we assessed heterogeneity through directionality analyses, post hoc testing, and subgroup analyses according to disorder-risk group, ascertainment paradigm, and task domain, as described above. Similarly, standard publication bias statistics (e.g., Egger’s test) are not applicable to ALE and other coordinate-based meta-analyses; findings of the present study are interpreted under the possibility of publication bias and selective reporting.

The statistical analysis was carried out in R version 4.4.1 (R Project for Statistical Computing) and Python version 3.12.6 (Python Software Foundation) from September 2025 to October 2026. In addition to the Python package Nilearn, fsleyes was used for brain activation displays.^13,14^

**eResults**

**eTable 3: Participant Characteristics by Study**

| **Study** | **High Risk (N)** | **Control (N)** | **Mean Age** | **% Female** | **Disorder Risk Group** | **Ascertainment Paradigm** |
| --- | --- | --- | --- | --- | --- | --- |
| **Allen 2010**^15^ | 15 | 15 | 26.3 | 43.33 | psychosis | PR |
| **Allen 2012**^16^ | 41 | 24 | 24.69 | 33.85 | psychosis | PR |
| **Allen 2015**^17^ | 33 | 27 | 22.78 | 38.33 | psychosis | PR |
| **Auday 2018**^18^ | 30 | 37 | 10.97 | 56.72 | anxiety | PR |
| **Bar-Haim 2009**^19^ | 16 | 16 | 16.22 | 46.88 | anxiety | PR |
| **Bernard 2018**^20^ | 20 | 21 | 21.16 | 43.9 | psychosis | PR |
| **Blackford 2011**^21^ | 18 | 15 | 23.0 | 63.64 | anxiety | PR |
| **Bonner-Jackson 2007**^22^ | 21 | 38 | 20.97 | 66.12 | psychosis | FHR |
| **Bourque 2017**^23^ | 27 | 135 | 14.34 | 65.45 | psychosis | PR |
| **Broome 2009**^24^ | 17 | 15 | 24.76 | 28.12 | psychosis | PR |
| **Broome 2010**^25^ | 17 | 15 | 24.76 |  | psychosis | PR |
| **Brüne 2011**^26^ | 10 | 26 | 27.88 | 33.33 | psychosis | PR |
| **Cao 2019**^27^ | 155 | 108 | 19.42 | 51.45 | psychosis | PR |
| **Carew 2013**^28^ | 16 | 16 | 21.4 |  | mood | FHR |
| **Chai 2015**^29^ | 36 | 14 | 11.24 | 48.0 | mood | FHR |
| **Chang 2017**^3^ | 50 | 29 | 13.54 | 44.2 | mood | PR; FHR |
| **Choi_GHR 2012**^4^ | 17 | 16 | 21.03 | 45.45 | psychosis | FHR |
| **Choi_UHR 2012**^4^ | 21 | 16 | 21.51 | 43.24 | psychosis | PR |
| **Christensen 2015**^30^ | 20 | 19 | 9.65 | 66.67 | anxiety | FHR |
| **Clauss 2011**^31^ | 18 | 19 | 23.42 | 59.38 | anxiety | PR |
| **Clauss 2014**^32^ | 17 | 17 | 21.75 | 60.0 | anxiety | PR |
| **Clauss 2016**^33^ | 18 | 19 | 9.65 | 54.3 | anxiety | PR |
| **Cole 2020**^34^ | 13 | 13 | 24.2 | 42.31 | psychosis | PR |
| **Colibazzi 2015**^35^ | 56 | 48 | 20.94 | 31.38 | psychosis | PR |
| **Collin 2021**^36^ | 15 | 19 | 9.43 | 58.82 | psychosis | FHR |
| **de Leeuw 2013**^37^ | 23 | 24 | 29.18 | 44.68 | psychosis | FHR |
| **Dedovic 2014**^38^ | 23 | 26 | 21.9 | 51.02 | mood | PR |
| **Dedovic 2016**^39^ | 22 | 26 | 21.9 | 50.0 | mood | PR |
| **Dodell-Feder 2014**^40^ | 19 | 18 | 26.82 | 75.68 | psychosis | FHR |
| **Falkenberg 2015**^41^ | 17 | 15 | 24.91 | 31.25 | psychosis | PR |
| **Falkenberg 2017**^42^ | 34 | 20 | 23.3 | 33.33 | psychosis | PR |
| **Fryer 2019**^43^ | 30 | 72 | 18.49 | 46.08 | psychosis | PR |
| **Fusar-Poli 2010a**^44^ | 15 | 15 | 24.77 | 43.33 | psychosis | PR |
| **Fusar-Poli 2010b**^45^ | 20 | 14 | 26.15 |  | psychosis | PR |
| **Fusar-Poli 2011a**^46^ | 15 | 15 | 24.77 | 43.33 | psychosis | PR |
| **Fusar-Poli 2011b**^47^ | 15 | 15 | 24.77 | 43.33 | psychosis | PR |
| **Garrett 2015**^48^ | 12 | 12 | 13.57 | 66.67 | mood | FHR |
| **Gawda 2016**^49^ | 10 | 7 | 26.12 | 52.94 | anxiety | PR |
| **Gisselgård 2018**^50^ | 41 | 37 | 16.79 | 56.26 | psychosis | PR |
| **Guyer 2014**^51^ | 18 | 21 | 17.89 | 56.41 | anxiety | PR |
| **Haut 2015**^52^ | 28 | 20 | 17.84 | 29.42 | psychosis | PR |
| **Hou 2022**^53^ | 21 | 23 | 19.96 | 34.09 | psychosis | PR |
| **Jacobson 2010**^54^ | 11 | 14 | 12.38 | 72.4 | psychosis | PR |
| **Jarcho 2016**^55^ | 30 | 23 | 11.08 | 45.28 | anxiety | PR |
| **Joormann 2012**^56^ | 20 | 27 | 11.81 | 100.0 | mood | FHR |
| **Juckel 2012**^57^ | 13 | 13 | 25.58 | 15.38 | psychosis | PR |
| **Karcher 2019a**^58^ | 21 | 20 | 18.32 | 70.72 | psychosis | PR |
| **Karcher 2019b**^59^ | 21 | 20 | 18.32 | 70.72 | psychosis | PR |
| **Karlsgodt 2014**^60^ | 20 | 19 | 17.33 | 30.77 | psychosis | PR |
| **Keshavan 2002**^61^ | 4 | 4 | 12.88 | 50.0 | psychosis | FHR |
| **Kim 2012**^62^ | 13 | 21 | 13.8 | 44.12 | mood | FHR |
| **Li 2007**^63^ | 15 | 15 | 22.8 | 53.33 | psychosis | FHR |
| **Li 2016**^64^ | 26 | 25 | 24.91 | 65.12 | psychosis | FHR |
| **Loeb 2018**^65^ | 30 | 39 | 19.74 | 44.93 | psychosis | FHR |
| **Macdonald 2016**^66^ | 16 | 21 | 22.0 | 48.65 | mood | FHR |
| **Mannie 2008**^67^ | 18 | 16 | 19.85 | 55.88 | mood | FHR |
| **Mannie 2010**^68^ | 17 | 15 | 18.25 | 78.12 | mood | FHR |
| **Mannie 2011**^69^ | 28 | 28 | 19.24 | 51.79 | mood | FHR |
| **Mannie 2014**^70^ | 62 | 59 | 18.9 | 61.16 | mood | FHR |
| **Meyhöfer 2015**^71^ | 12 | 19 | 23.61 | 58.05 | psychosis | PR |
| **Michielse 2019**^72^ | 47 | 40 | 21.68 | 83.62 | psychosis | PR |
| **Millman 2020**^73^ | 22 | 19 | 17.62 | 46.3 | psychosis | PR |
| **Millman 2022**^74^ | 26 | 19 | 17.78 | 48.53 | psychosis | PR |
| **Modinos 2010a**^75^ | 17 | 17 | 20.4 | 58.82 | psychosis | PR |
| **Modinos 2010b**^76^ | 18 | 18 | 20.4 | 44.44 | psychosis | PR |
| **Morgan 2019**^77^ | 25 | 31 | 6.8 | 55.57 | mood | FHR |
| **Mori 2016**^78^ | 15 | 15 | 18.8 | 53.33 | mood | PR |
| **Niendam 2014**^79^ | 25 | 35 | 17.29 | 45.17 | psychosis | PR |
| **Nook 2018**^80^ | 21 | 19 | 26.7 | 70.0 | psychosis | FHR |
| **Olino 2015**^81^ | 10 | 23 | 12.98 | 57.4 | mood | FHR |
| **Park 2016**^82^ | 20 | 17 | 23.51 | 59.46 | psychosis | FHR |
| **Park 2020**^83^ | 22 | 28 | 21.69 | 37.64 | psychosis | PR |
| **Pauly 2010**^84^ | 12 | 12 | 24.34 | 16.67 | psychosis | PR |
| **Pilhatsch 2014**^85^ | 28 | 136 | 14.56 | 50.61 | mood | FHR |
| **Premkumar 2013**^86^ | 12 | 12 | 29.29 | 79.17 | psychosis | PR |
| **Pulkkinen 2015**^87^ | 51 | 52 | 22.35 | 61.15 | psychosis | FHR |
| **Roberts 2013**^88^ | 47 | 49 | 23.89 | 59.38 | mood | FHR |
| **Rzepa 2017**^89^ | 16 | 17 | 16.43 | 69.7 | mood | PR |
| **Schmidt 2017**^90^ | 23 | 13 | 22.58 | 38.89 | psychosis | PR |
| **Seiferth 2008**^91^ | 12 | 12 | 24.5 | 16.67 | psychosis | PR |
| **Shechner 2018**^92^ | 22 | 28 | 20.5 | 48.0 | anxiety | PR |
| **Singh 2014**^93^ | 20 | 25 | 12.19 | 62.22 | mood | FHR |
| **Smieskova 2015**^94^ | 34 | 19 | 25.09 | 32.08 | psychosis | PR |
| **Stolz 2012**^95^ | 16 | 28 | 25.48 | 65.91 | psychosis | FHR |
| **Thermenos 2011**^96^ | 10 | 10 | 17.75 | 50.0 | mood | FHR |
| **Thermenos 2013**^97^ | 43 | 32 | 24.94 | 65.88 | psychosis | FHR |
| **Thermenos 2016**^98^ | 37 | 34 | 19.79 | 53.25 | psychosis | PR |
| **Tseng 2015**^99^ | 13 | 37 | 14.44 | 52.04 | mood | FHR |
| **Valli 2016**^100^ | 25 | 25 | 24.48 | 36.0 | psychosis | PR |
| **van Buuren 2011**^101^ | 24 | 25 | 28.43 | 69.39 | psychosis | FHR |
| **van der Velde 2015**^102^ | 15 | 16 | 22.58 | 48.55 | psychosis | PR |
| **van Gool 2022**^103^ | 17 | 20 | 9.44 | 57.6 | psychosis | FHR |
| **Venkatasubramanian 2010**^104^ | 17 | 16 | 24.81 | 15.15 | psychosis | FHR |
| **Whyte 2006**^105^ | 68 | 21 | 26.19 | 50.56 | psychosis | FHR |
| **Wiggins 2017**^106^ | 27 | 19 | 7.52 | 61.09 | mood | FHR |
| **Wilson 2019**^107^ | 17 | 19 | 23.99 | 49.99 | psychosis | PR |
| **Winton-Brown 2017**^108^ | 29 | 32 | 22.49 | 49.18 | psychosis | PR |
| **Wotruba 2014**^109^ | 21 | 24 | 24.14 | 38.84 | psychosis | PR |
| **Yaakub 2013**^110^ | 60 | 38 | 22.0 | 36.74 | psychosis | PR |
| **Young 2016**^111^ | 30 | 60 | 30.4 | 54.43 | mood | FHR |
| **Yun 2022**^112^ | 21 | 27 | 24.51 | 43.21 | mood | PR |
| **Zhang 2022**^113^ | 42 | 32 | 21.89 | 68.91 | mood | PR |

Note: FHR = familial high-risk; PR = phenotypic risk

**eTable 4: Task Characteristics by Study**

| **Study** | **Task Details** | **Task Type** |
| --- | --- | --- |
| **Allen 2010**^15^ | Hayling Sentence Completion task | cognitive |
| **Allen 2012**^16^ | verbal fluency task | cognitive |
| **Allen 2015**^17^ | verbal fluency task | cognitive |
| **Auday 2018**^18^ | motion-face masked dot-probe task | affective |
| **Bar-Haim 2009**^19^ | reward-contingency task | reward |
| **Bernard 2018**^20^ | second-order rule learning | cognitive |
| **Blackford 2011**^21^ | face recognition task | cognitive |
| **Bonner-Jackson 2007**^22^ | word encoding task | cognitive |
| **Bourque 2017**^23^ | (1) stop signal task; (2) faces task; (3) monetary incentive delay | (1) cognitive; (2) affective; (3) reward |
| **Broome 2009**^24^ | (1) N-back task; (2) verbal fluency task | (1) cognitive; (2) cognitive |
| **Broome 2010**^25^ | random movement generation task | cognitive |
| **Brüne 2011**^26^ | theory of mind task | cognitive |
| **Cao 2019**^27^ | paired-associate episodic memory task | cognitive |
| **Carew 2013**^28^ | thought suppression task | cognitive |
| **Chai 2015**^29^ | face-match task | affective |
| **Chang 2017**^3^ | facial expressions task | affective |
| **Choi_GHR 2012**^4^ | spatial delayed-response task | cognitive |
| **Choi_UHR 2012**^4^ | spatial delayed-response task | cognitive |
| **Christensen 2015**^30^ | dot-probe attention-orienting task | affective |
| **Clauss 2011**^31^ | expected compared to unexpected fear faces task | affective |
| **Clauss 2014**^32^ | anticipation task | affective |
| **Clauss 2016**^33^ | cued anticipation task | affective |
| **Cole 2020**^34^ | probabilistic learning task | reward |
| **Colibazzi 2015**^35^ | Simon task | cognitive |
| **Collin 2021**^36^ | self-reference task | cognitive |
| **de Leeuw 2013**^37^ | Sternberg working memory task | cognitive |
| **Dedovic 2014**^38^ | modified Montreal imaging stress task | cognitive |
| **Dedovic 2016**^39^ | dot-probe task | affective |
| **Dodell-Feder 2014**^40^ | person-description task | cognitive |
| **Falkenberg 2015**^41^ | N-back task | cognitive |
| **Falkenberg 2017**^42^ | delayed matching to sample task | cognitive |
| **Fryer 2019**^43^ | Go/NoGo task | cognitive |
| **Fusar-Poli 2010a**^44^ | visuospatial working memory task | cognitive |
| **Fusar-Poli 2010b**^45^ | N-back task | cognitive |
| **Fusar-Poli 2011a**^46^ | N-back task | cognitive |
| **Fusar-Poli 2011b**^47^ | verbal fluency task | cognitive |
| **Garrett 2015**^48^ | facial expression task | affective |
| **Gawda 2016**^49^ | verbal fluency task | cognitive |
| **Gisselgård 2018**^50^ | N-back task | cognitive |
| **Guyer 2014**^51^ | social evaluation task | affective |
| **Haut 2015**^52^ | remember-know task | cognitive |
| **Hou 2022**^53^ | facial emotion recognition task | affective |
| **Jacobson 2010**^54^ | Go/NoGo task | cognitive |
| **Jarcho 2016**^55^ | peer-interaction task | affective |
| **Joormann 2012**^56^ | mood-repair task | affective |
| **Juckel 2012**^57^ | monetary incentive delay task | reward |
| **Karcher 2019a**^58^ | Weather Prediction Task | reward |
| **Karcher 2019b**^59^ | reversal learning task | reward |
| **Karlsgodt 2014**^60^ | Sternberg-style verbal working memory task | cognitive |
| **Keshavan 2002**^61^ | saccade task | cognitive |
| **Kim 2012**^62^ | change task | cognitive |
| **Li 2007**^63^ | visual word discrimination task | cognitive |
| **Li 2016**^64^ | verbal- versus spatial-1-back working memory task | cognitive |
| **Loeb 2018**^65^ | N-back task | cognitive |
| **Macdonald 2016**^66^ | narrative task | affective |
| **Mannie 2008**^67^ | emotional counting Stroop task | affective |
| **Mannie 2010**^68^ | N-back task | cognitive |
| **Mannie 2011**^69^ | emotional face matching task | affective |
| **Mannie 2014**^70^ | memory encoding task | cognitive |
| **Meyhöfer 2015**^71^ | Smooth pursuit eye movement task | cognitive |
| **Michielse 2019**^72^ | monetary incentive delay task | reward |
| **Millman 2020**^73^ | card betting task | reward |
| **Millman 2022**^74^ | monetary incentive delay task | reward |
| **Modinos 2010a**^75^ | viewing images task | affective |
| **Modinos 2010b**^76^ | theory of mind task | cognitive |
| **Morgan 2019**^77^ | faces task | affective |
| **Mori 2016**^78^ | monetary incentive delay task | reward |
| **Niendam 2014**^79^ | AX continuous performance task | cognitive |
| **Nook 2018**^80^ | expressed emotion task | affective |
| **Olino 2015**^81^ | chatroom interact task | cognitive |
| **Park 2016**^82^ | facial emotion processing task | affective |
| **Park 2020**^83^ | self-referential processing task | cognitive |
| **Pauly 2010**^84^ | facial emotion processing task | cognitive |
| **Pilhatsch 2014**^85^ | perceptual discrimination task | affective |
| **Premkumar 2013**^86^ | response to critical and positive comments from relatives task | affective |
| **Pulkkinen 2015**^87^ | dynamic happy and fearful faces viewing task | affective |
| **Roberts 2013**^88^ | facial emotion go/no-go task | affective |
| **Rzepa 2017**^89^ | processing adverse and rewarding stimuli task | reward |
| **Schmidt 2017**^90^ | salience attribution task | reward |
| **Seiferth 2008**^91^ | emotion discrimination task | affective |
| **Shechner 2018**^92^ | extinction recall task | affective |
| **Singh 2014**^93^ | monetary incentive delay task | reward |
| **Smieskova 2015**^94^ | salience attribution task | reward |
| **Stolz 2012**^95^ | visual episodic memory encoding and retrieval task | cognitive |
| **Thermenos 2011**^96^ | N-back task | cognitive |
| **Thermenos 2013**^97^ | lexical decision task | cognitive |
| **Thermenos 2016**^98^ | N-back task | cognitive |
| **Tseng 2015**^99^ | encoding faces task | affective |
| **Valli 2016**^100^ | verbal memory task | cognitive |
| **van Buuren 2011**^101^ | emotion processing task | affective |
| **van der Velde 2015**^102^ | emotion regulation task | affective |
| **van Gool 2022**^103^ | N-back task | cognitive |
| **Venkatasubramanian 2010**^104^ | facial emotion recognition task | affective |
| **Whyte 2006**^105^ | (1) word classification task; (2) retrieval task | (1) cognitive; (2) cognitive |
| **Wiggins 2017**^106^ | monetary incentive delay task | reward |
| **Wilson 2019**^107^ | monetary incentive delay task | reward |
| **Winton-Brown 2017**^108^ | salience integration task | reward |
| **Wotruba 2014**^109^ | monetary incentive delay task | reward |
| **Yaakub 2013**^110^ | verbal working memory task | cognitive |
| **Young 2016**^111^ | autobiographical memory task | affective |
| **Yun 2022**^112^ | Simon task | reward |
| **Zhang 2022**^113^ | passive viewing task | affective |

**eTable 5: Quality Assessment**

| **Study** | **Rating 1** | **Rating 2** |
| --- | --- | --- |
| **Allen 2010**^15^ | 10.0 | 10.0 |
| **Allen 2012**^16^ | 10.0 | 9.5 |
| **Allen 2015**^17^ | 10.0 | 10.0 |
| **Auday 2018**^18^ | 9.5 | 10.0 |
| **Bar-Haim 2009**^19^ | 10.0 | 10.0 |
| **Bernard 2018**^20^ | 10.0 | 9.0 |
| **Blackford 2011**^21^ | 10.0 | 10.0 |
| **Bonner-Jackson 2007**^22^ | 10.0 | 10.0 |
| **Bourque 2017**^23^ | 10.0 | 9.0 |
| **Broome 2009**^24^ | 9.5 | 10.0 |
| **Broome 2010**^25^ | 10.0 | 10.0 |
| **Brüne 2011**^26^ | 9.5 | 10.0 |
| **Cao 2019**^27^ | 10.0 | 10.0 |
| **Carew 2013**^28^ | 10.0 | 10.0 |
| **Chai 2015**^29^ | 10.0 | 9.5 |
| **Chang 2017**^3^ | 10.0 | 10.0 |
| **Choi 2012**^4^ | 10.0 | 10.0 |
| **Christensen 2015**^30^ | 10.0 | 9.5 |
| **Clauss 2011**^31^ | 10.0 | 10.0 |
| **Clauss 2014**^32^ | 10.0 | 10.0 |
| **Clauss 2016**^33^ | 10.0 | 10.0 |
| **Cole 2020**^34^ | 10.0 | 9.5 |
| **Colibazzi 2015**^35^ | 10.0 | 9.5 |
| **Collin 2021**^36^ | 10.0 | 9.5 |
| **de Leeuw 2013**^37^ | 10.0 | 10.0 |
| **Dedovic 2014**^38^ | 10.0 | 9.5 |
| **Dedovic 2016**^39^ | 10.0 | 9.0 |
| **Dodell-Feder 2014**^40^ | 10.0 | 9.5 |
| **Falkenberg 2015**^41^ | 10.0 | 10.0 |
| **Falkenberg 2017**^42^ | 9.5 | 9.5 |
| **Fryer 2019**^43^ | 10.0 | 10.0 |
| **Fusar-Poli 2010a**^44^ | 10.0 | 10.0 |
| **Fusar-Poli 2010b**^45^ | 10.0 | 10.0 |
| **Fusar-Poli 2011a**^46^ | 10.0 | 10.0 |
| **Fusar-Poli 2011b**^47^ | 10.0 | 10.0 |
| **Garrett 2015**^48^ | 10.0 | 10.0 |
| **Gawda 2016**^49^ | 9.5 | 9.0 |
| **Gisselgård 2018**^50^ | 10.0 | 9.5 |
| **Guyer 2014**^51^ | 10.0 | 9.5 |
| **Haut 2015**^52^ | 9.5 | 9.5 |
| **Hou 2022**^53^ | 10.0 | 10.0 |
| **Jacobson 2010**^54^ | 9.5 | 10.0 |
| **Jarcho 2016**^55^ | 9.0 | 10.0 |
| **Joormann 2012**^56^ | 10.0 | 9.0 |
| **Juckel 2012**^57^ | 9.5 | 9.0 |
| **Karcher 2019a**^58^ | 10.0 | 10.0 |
| **Karcher 2019b**^59^ | 10.0 | 9.0 |
| **Karlsgodt 2014**^60^ | 10.0 | 9.5 |
| **Keshavan 2002**^61^ | 9.0 | 8.0 |
| **Kim 2012**^62^ | 10.0 | 10.0 |
| **Li 2007**^63^ | 10.0 | 10.0 |
| **Li 2016**^64^ | 10.0 | 10.0 |
| **Loeb 2018**^65^ | 10.0 | 10.0 |
| **Macdonald 2016**^66^ | 10.0 | 9.0 |
| **Mannie 2008**^67^ | 10.0 | 9.5 |
| **Mannie 2010**^68^ | 10.0 | 9.5 |
| **Mannie 2011**^69^ | 10.0 | 9.0 |
| **Mannie 2014**^70^ | 10.0 | 9.5 |
| **Meyhöfer 2015**^71^ | 10.0 | 10.0 |
| **Michielse 2019**^72^ | 10.0 | 10.0 |
| **Millman 2020**^73^ | 10.0 | 10.0 |
| **Millman 2022**^74^ | 10.0 | 10.0 |
| **Modinos 2010a**^75^ | 10.0 | 9.5 |
| **Modinos 2010b**^76^ | 10.0 | 9.5 |
| **Morgan 2019**^77^ | 9.5 | 9.0 |
| **Mori 2016**^78^ | 10.0 | 9.5 |
| **Niendam 2014**^79^ | 9.5 | 10.0 |
| **Nook 2018**^80^ | 9.0 | 10.0 |
| **Olino 2015**^81^ | 8.5 | 8.5 |
| **Park 2016**^82^ | 10.0 | 10.0 |
| **Park 2020**^83^ | 9.5 | 9.5 |
| **Pauly 2010**^84^ | 10.0 | 10.0 |
| **Pilhatsch 2014**^85^ | 10.0 | 10.0 |
| **Premkumar 2013**^86^ | 10.0 | 10.0 |
| **Pulkkinen 2015**^87^ | 10.0 | 10.0 |
| **Roberts 2013**^88^ | 10.0 | 9.5 |
| **Rzepa 2017**^89^ | 10.0 | 9.0 |
| **Schmidt 2017**^90^ | 10.0 | 9.5 |
| **Seiferth 2008**^91^ | 10.0 | 10.0 |
| **Shechner 2018**^92^ | 9.5 | 10.0 |
| **Singh 2014**^93^ | 10.0 | 10.0 |
| **Smieskova 2015**^94^ | 10.0 | 10.0 |
| **Stolz 2012**^95^ | 9.5 | 9.5 |
| **Thermenos 2011**^96^ | 9.5 | 10.0 |
| **Thermenos 2013**^97^ | 9.5 | 10.0 |
| **Thermenos 2016**^98^ | 10.0 | 10.0 |
| **Tseng 2015**^99^ | 10.0 | 9.0 |
| **Valli 2016**^100^ | 10.0 | 10.0 |
| **van Buuren 2011**^101^ | 9.5 | 9.0 |
| **van der Velde 2015**^102^ | 10.0 | 9.0 |
| **van Gool 2022**^103^ | 10.0 | 9.5 |
| **Venkatasubramanian 2010**^104^ | 9.5 | 9.5 |
| **Whyte 2006**^105^ | 10.0 | 10.0 |
| **Wiggins 2017**^106^ | 9.5 | 9.5 |
| **Wilson 2019**^107^ | 10.0 | 10.0 |
| **Winton-Brown 2017**^108^ | 10.0 | 10.0 |
| **Wotruba 2014**^109^ | 10.0 | 10.0 |
| **Yaakub 2013**^110^ | 10.0 | 10.0 |
| **Young 2016**^111^ | 10.0 | 8.5 |
| **Yun 2022**^112^ | 10.0 | 10.0 |
| **Zhang 2022**^113^ | 9.5 | 10.0 |

Convergence in Brain Activation across Disorder-Risk Groups

*Overall Meta-Analysis (cFWE):*

All ascertainment paradigms and task domains contributed to the cluster (**eFigure 2**). Additionally, the sample size used for Gaussian kernel weighting (smaller sample size across control and risk groups) was similar across contributing samples (10-32; Figure 2B) and the relative contribution of each sample was similar as well (0.13-6.39% for 28/30 and ~10% for 2/30 studies; Figure 2B). Post-hoc statistical testing for systematic differences between samples contributing to this cluster (30 studies) and those not contributing (72 studies) revealed no differences in mean age, percent female, disorder-risk group, ascertainment paradigm distribution, and task domain distribution (all p>0.3). Data on sex was not available for 1 sample contributing to the cluster and for 2 samples not contributing. One sample’s risk group was defined through both ascertainment paradigms, and one sample had one task from each task domain. These were excluded from the post-hoc testing for ascertainment paradigm and task domain, respectively.

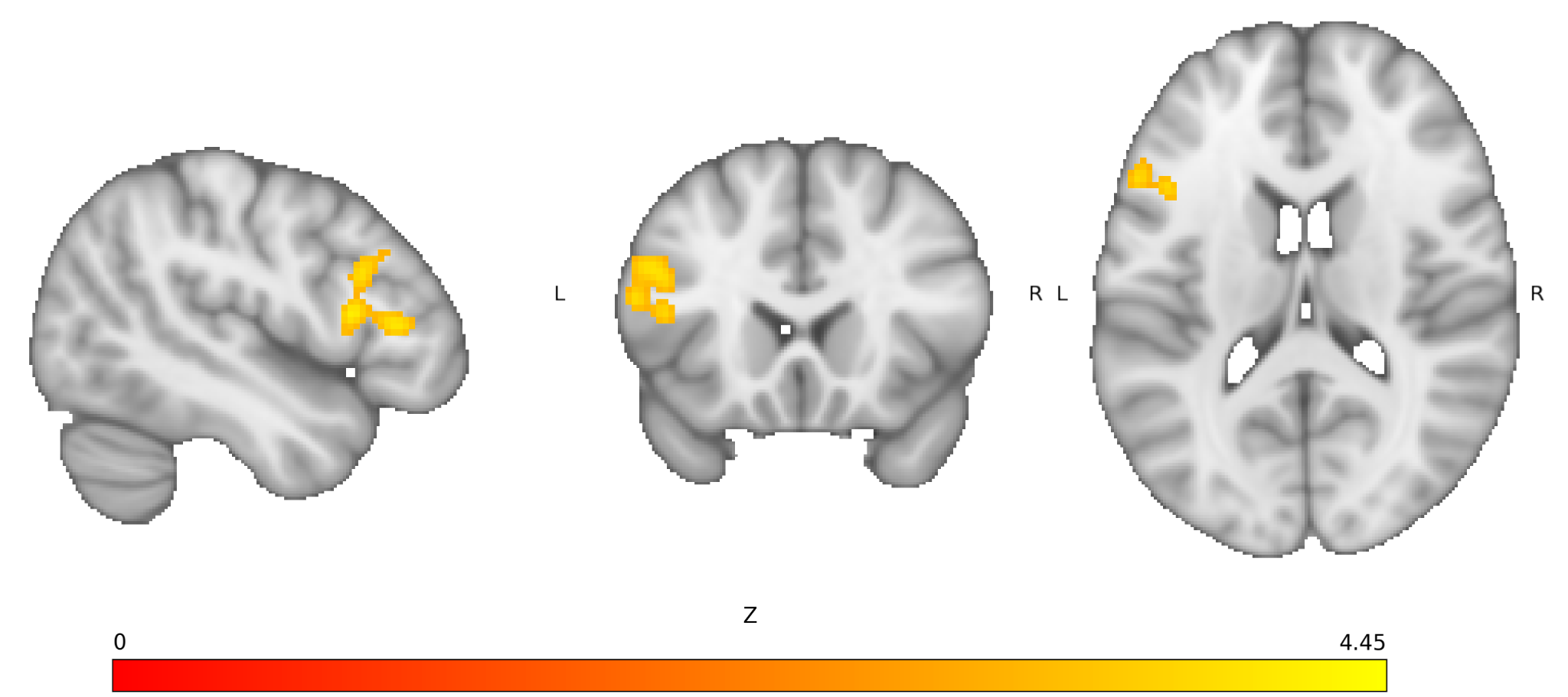
**eFigure 1: Brain Activation Convergence (p < .05, cFWE) in the Overall Meta-Analysis across All Samples.**

The cluster is in the left inferior and middle frontal gyrus left inferior and middle frontal gyrus, spanning the ventrolateral prefrontal cortex (vlPFC) and dorsolateral prefrontal cortex (dlPFC; k=319 voxels; center: x=-46, y=20, z=16).

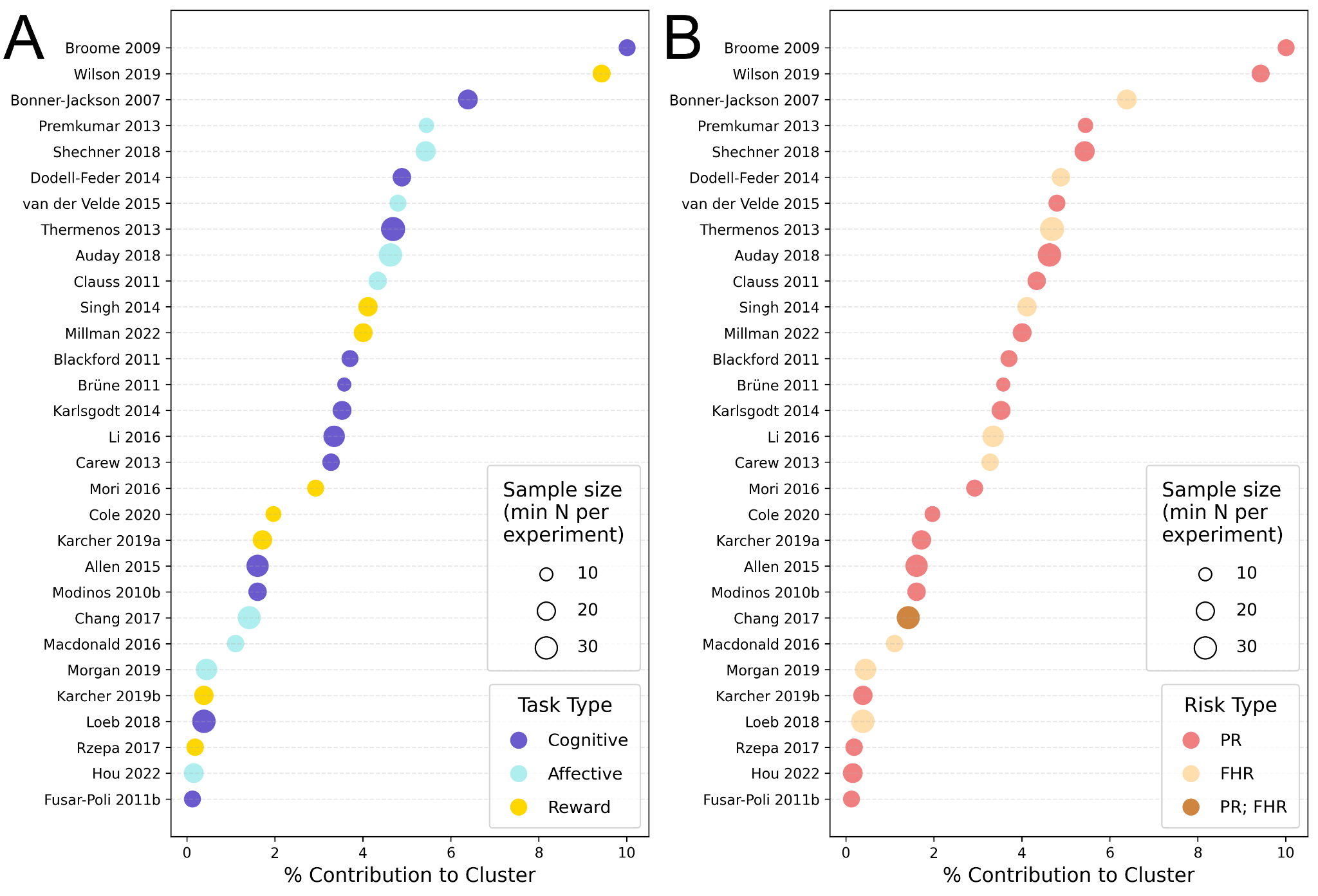
 **eFigure 2: All (A) Task Domains and (B) Ascertainment Paradigms Contributed to Brain Activation Convergence in the Overall Meta-Analysis across All Samples**

Sample size here referred to the smaller size between the control and risk sample in each study, which was used for Gaussian kernel weighting in ALE. PR = phenotypic risk; FHR = familial high-risk.

*Directional Meta-Analysis (cFWE):*

Comparing high-risk samples > control samples showed that overall findings were driven by greater activation in the high-risk group compared to control (p < .05, FWE, **eFigure 3**). A second cluster was also identified in the frontal pole. There were no significant findings for control > risk (p > 0.05, FWE).

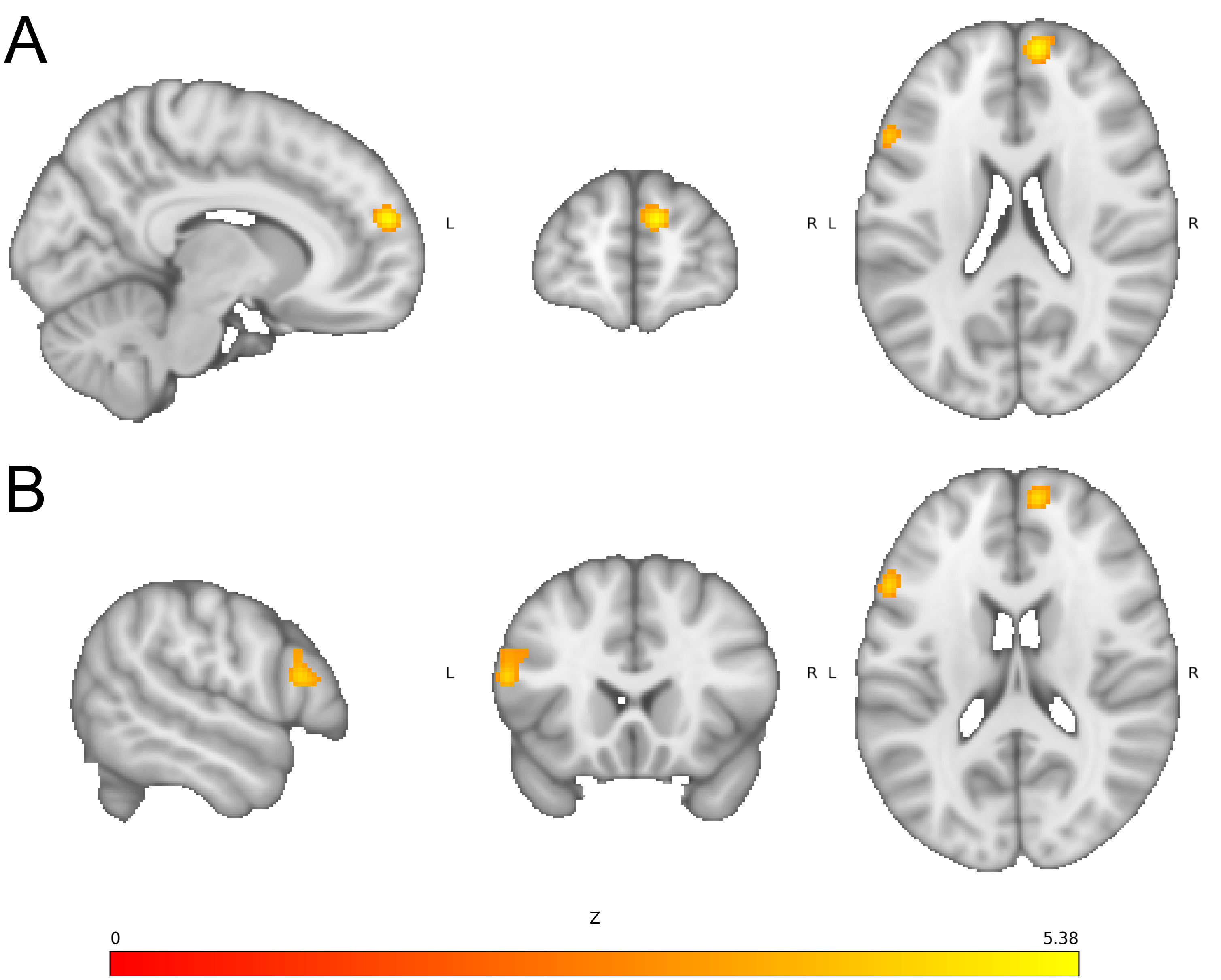

**eFigure 3: Brain Activation for High Risk > Control in the Overall Meta-Analysis Across All Samples (p <.05, cFWE).** A) Greater activation in the frontal pole/dorsomedial prefrontal cortex and B) greater activation in an area of the left dorsolateral prefrontal cortex largely overlapping with the overall meta-analysis (**Figure 2A, eFigure 1**).

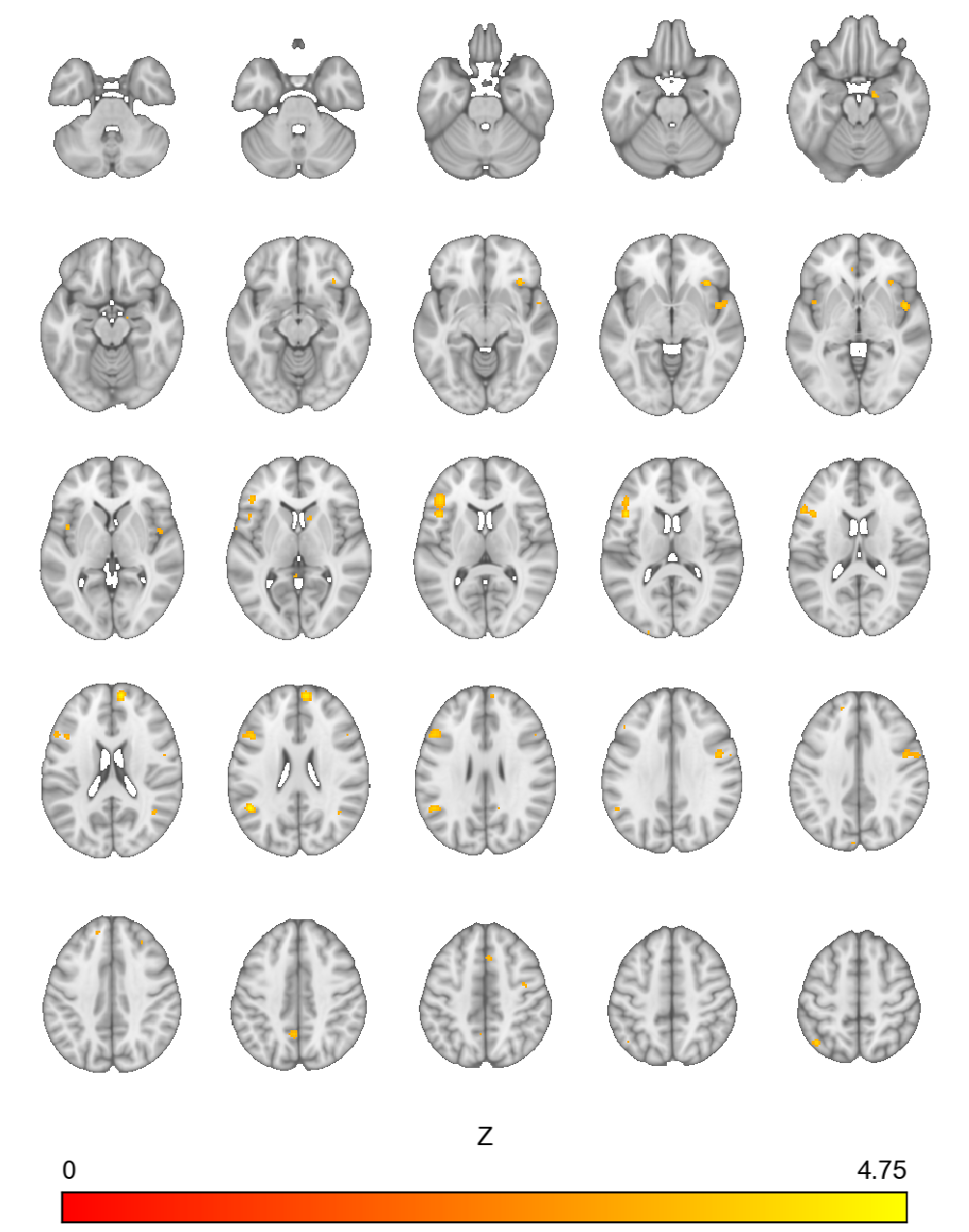

**eFigure 4: Brain Activation Convergence (p <.001, not cFWE corrected) in the Overall Meta-Analysis across All Samples.** Axial brain slices illustrate the spatial distribution of significant activation differences across the brain, with clusters observed in bilateral frontal, temporal, parietal, and occipital regions, right amygdala, and right caudate. Warm colors indicate increasing Z statistics (red to yellow; maximum displayed Z = 4.75). Areas of focal convergence across were identified at an uncorrected voxel-level p <.001.

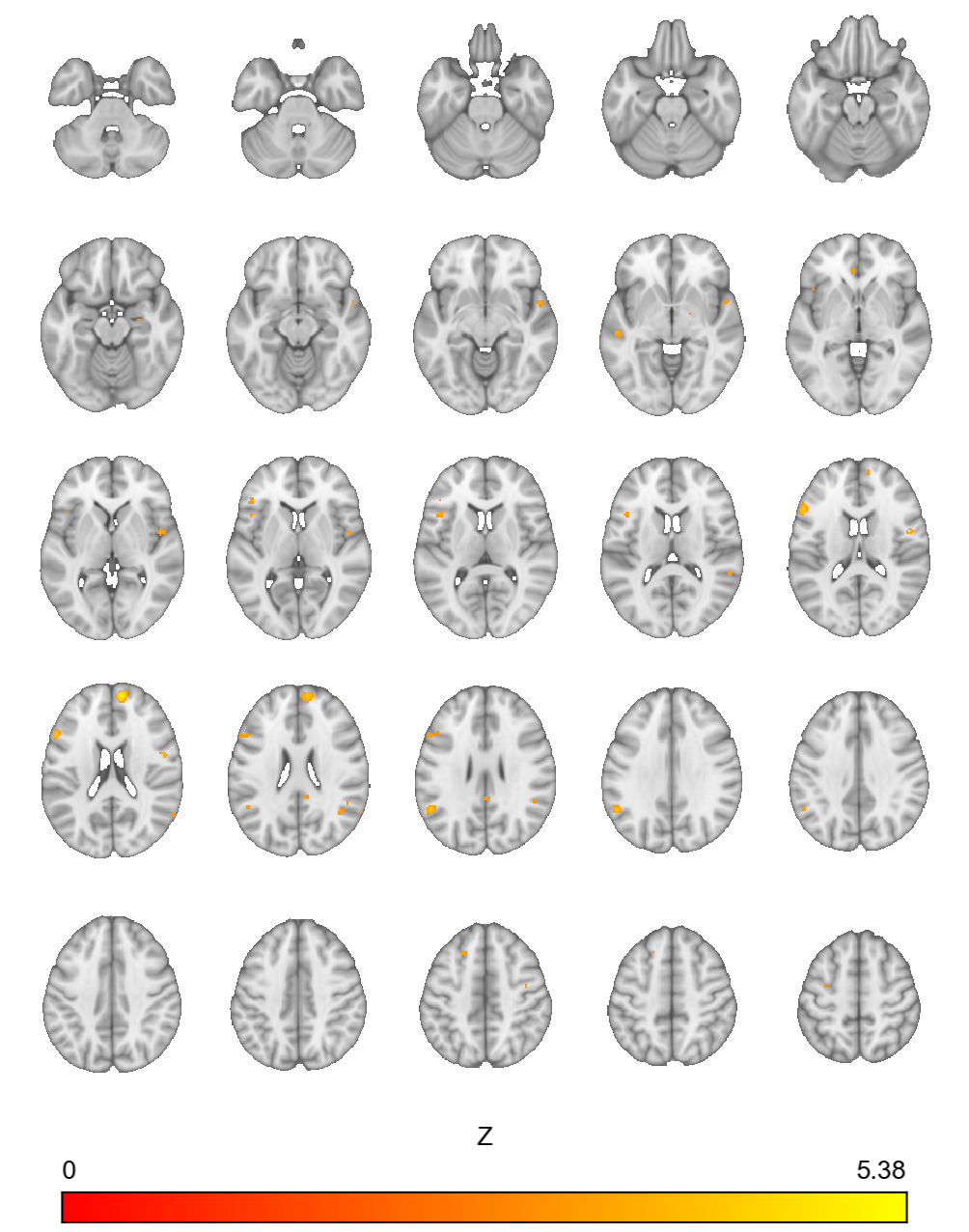

**eFigure 5: Brain Activation Convergence (p <.001, uncorrected) for High Risk > Control in the Overall Meta-Analysis Across All Samples.** Axial brain slices demonstrate clusters distributed across bilateral frontal, temporal, and parietal regions, including medial frontal cortex. Cortical regions largely overlap with overall findings in eFigure 4. Warm colors indicate increasing Z statistics, with a maximum displayed Z value of 5.38.

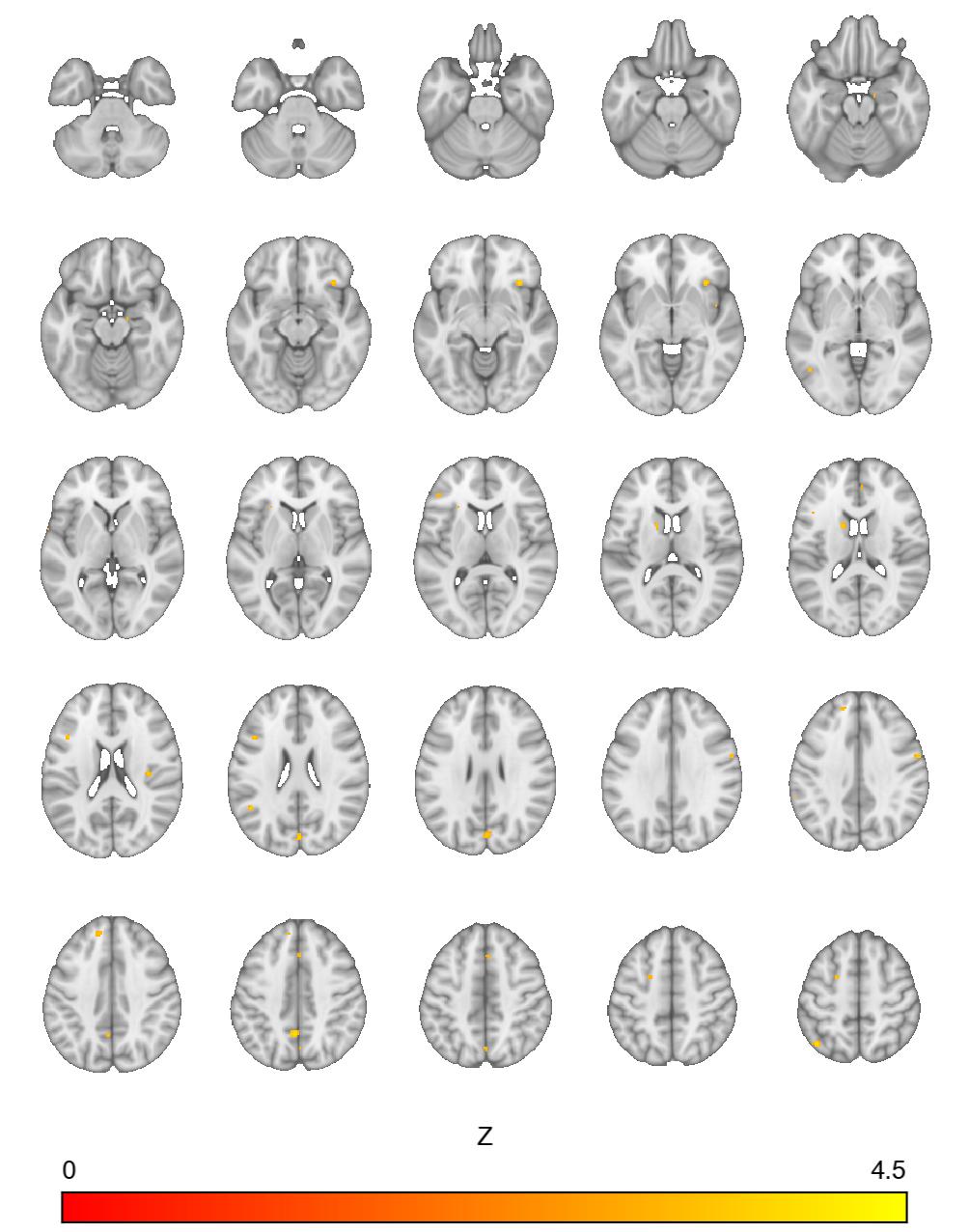

**eFigure 6. Brain Activation Convergence (p <.001, uncorrected) for Control > High Risk in the Overall Meta-Analysis Across All Samples.** Axial brain slices demonstrate scattered clusters in right insula, right hippocampus, frontal, temporal, and parietal regions, with additional clusters in posterior cortical regions, including posterior cingulate cortex. Warm colors indicate increasing Z statistics, with a maximum displayed Z value of 4.50.

Brain Activation Convergence by Disorder-Risk Group, Ascertainment Paradigm, and Task Domain

*Psychosis Risk Sample (cFWE):*

For the two clusters which included portions of the lateral PFC, samples which contributed to these findings spanned both psychosis-risk ascertainment paradigms (PR, FHR) and all three task domains (cognitive, reward, and affective). For cluster 2, which included the right anterior insula and OFC, only cognitive and reward tasks, and only PR samples contributed (**Figure 3A, eFigures 7-10**).

For all three clusters, the sample size used for Gaussian kernel weighting was consistent across contributing samples with two outliers (cluster 1: 12-20 for 9/10 and 4 for 1/10 samples, cluster 2: 12-40 for all 9 samples, cluster 3: 10-32 for 18/19 and 108 for 1/19 samples; **eFigures 8-10**). The percent contribution of each sample was consistent as well (cluster 1: 8.74-18.01% for 8/10 and under 4% for 2/10 samples, cluster 2: 11.38-18.43% for 6/9 and under 3% for 3/9 samples, cluster 3: 0.11-8.23% for 17/19 and ~17% for 2/19 samples; **eFigures 8-10).** For all three clusters, no statistically significant differences between included and excluded samples were found in terms of mean age, percent female, ascertainment paradigm distribution, or task domain distribution. Data on sex were not available for two samples, and the sample containing all three task domains was excluded when testing differences in task distribution.

###### eFigure 7: Brain Activation Convergence (p <.05, cFWE) in the Psychotic Disorder Risk Meta-Analysis

(A) Cluster 1 is located in the right precentral gyrus (center: x=48, y=0, z=34).

(B) Cluster 2 is in the right anterior insula and orbitofrontal cortex (center: x=34, y=24, z=-6).

(C) Cluster 3 is in the left inferior frontal gyrus (center: x=-46, y=18, z=12).

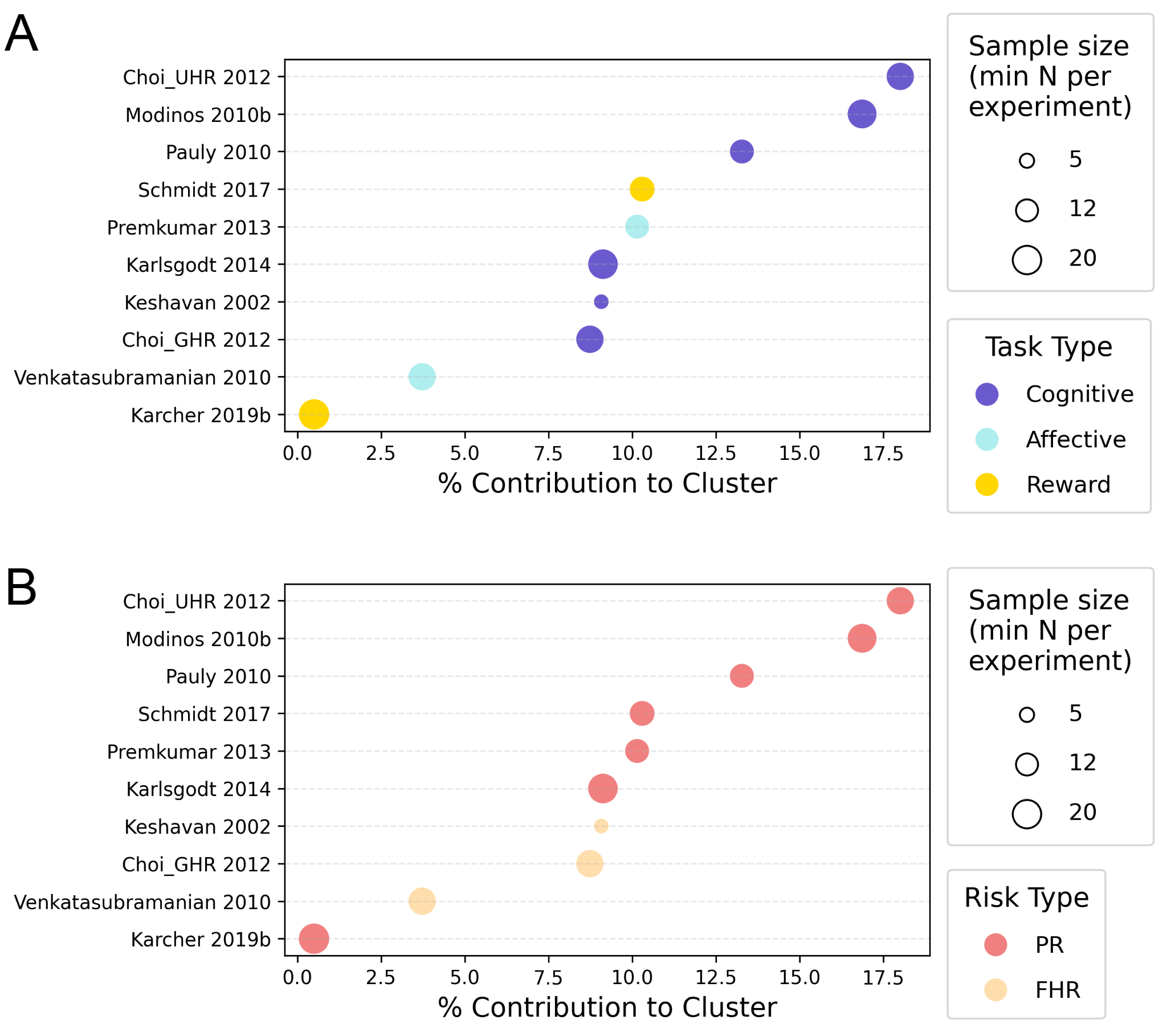

###### eFigure 8: All (A) Task Domains and (B) Ascertainment Paradigms Contributed to Brain Activation Convergence in Cluster 1 in the Psychotic Disorder Risk Meta-Analysis

Sample size here referred to the smaller size across the control and risk sample in each study, which was used for Gaussian kernel weighting in ALE. PR = phenotypic risk; FHR = familial high-risk.

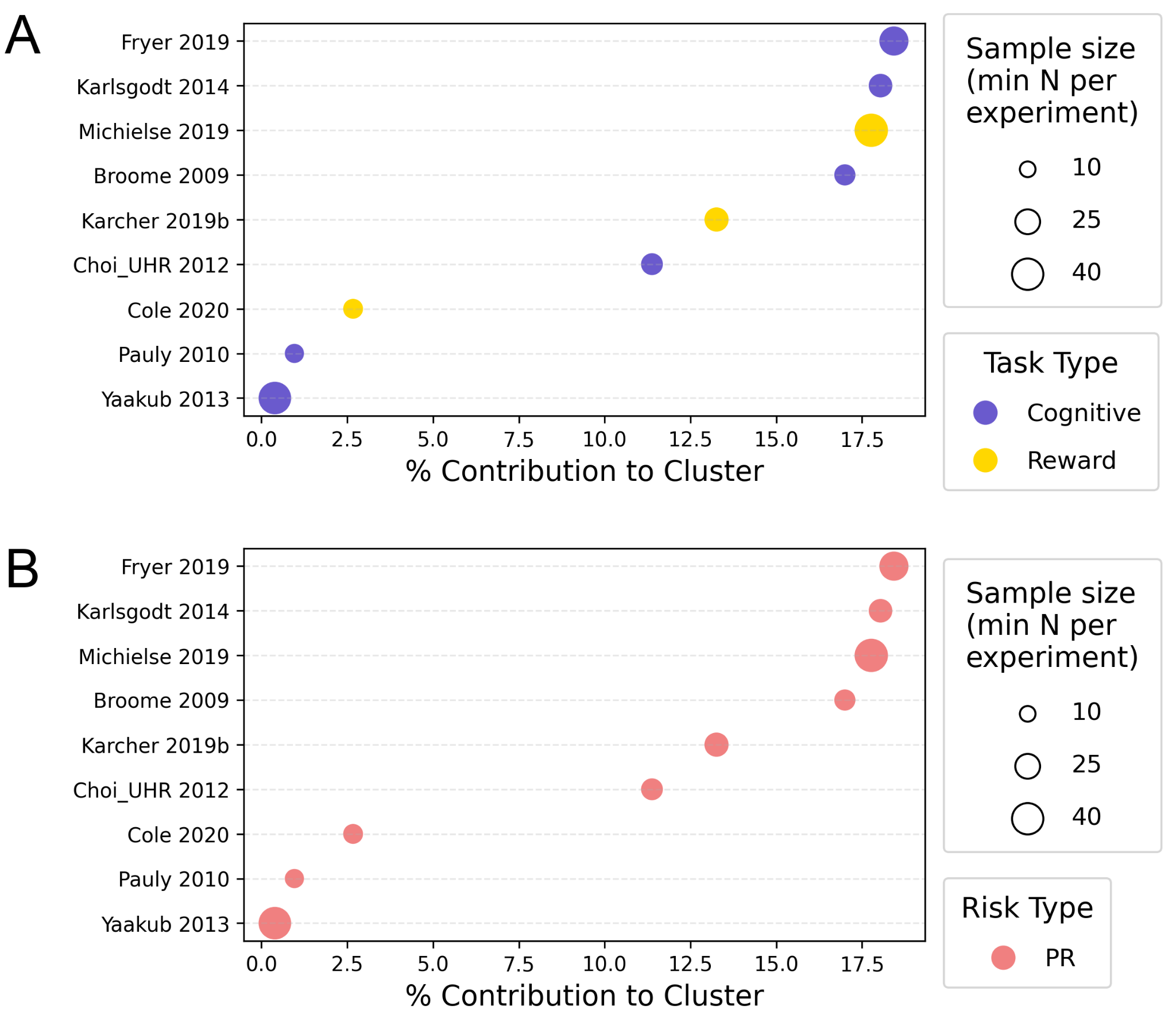

###### eFigure 9: (A) Cognitive and Reward Tasks and (B) Symptom and Trait-Based Risk Samples Contributed to Brain Activation Convergence in Cluster 2 in the Psychotic Disorder Risk Meta-Analysis

Sample size here referred to the smaller size across the control and risk sample in each study, which was used for Gaussian kernel weighting in ALE. PR = phenotypic risk; FHR = familial high-risk.

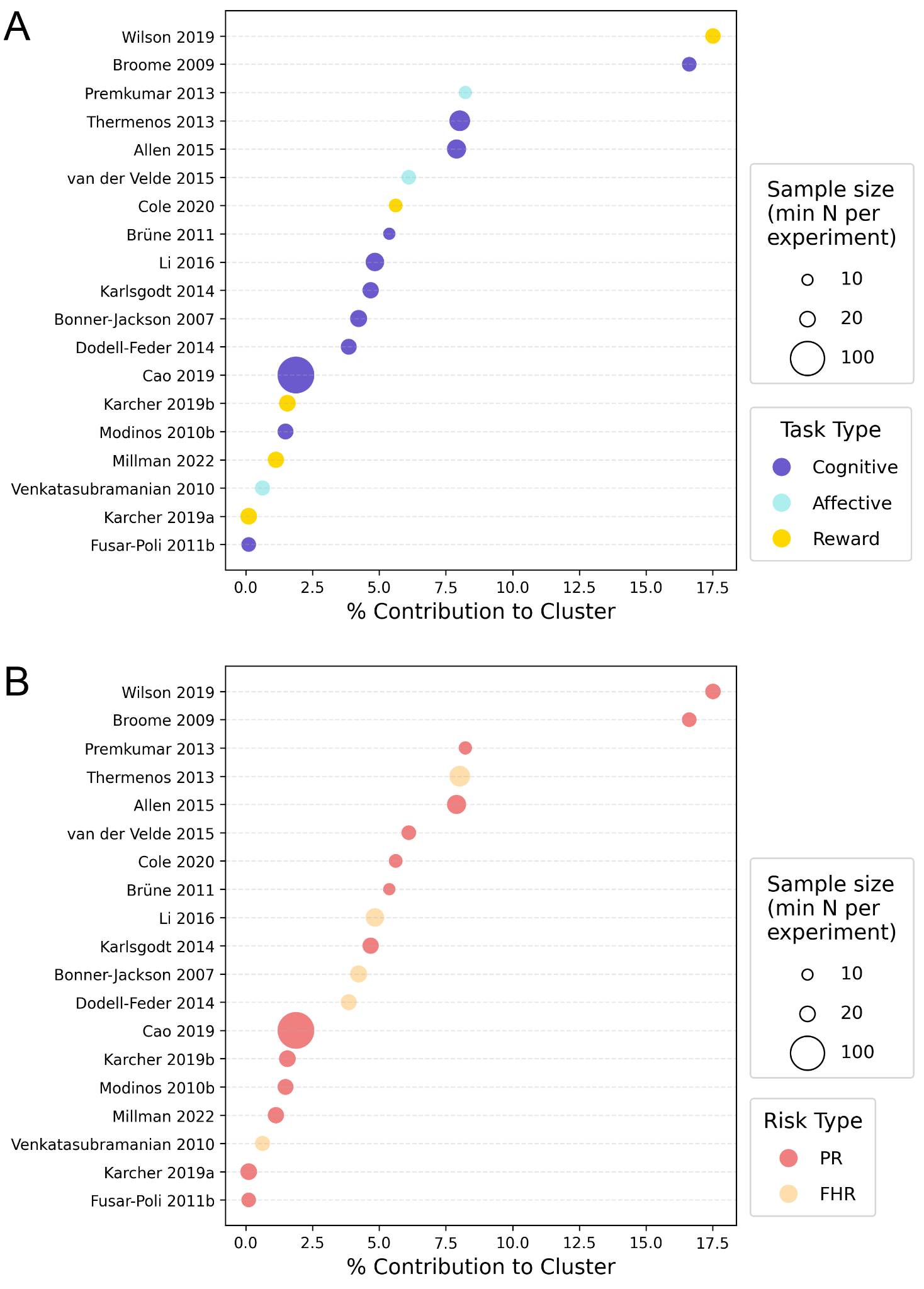

###### eFigure 10: All (A) Task Domains and (B) Ascertainment Paradigms Contributed to Brain Activation Convergence in Cluster 3 in the Psychotic Disorder Risk Meta-Analysis

Sample size here referred to the smaller size across the control and risk sample in each study, which was used for Gaussian kernel weighting in ALE. PR = phenotypic risk; FHR = familial high-risk.

*FHR Sample (cFWE):*

The combined FHR sample (N=39) had one statistically significant cluster (**Figure 3B, eFigure 11**): right frontal pole (k = 101; 6/39 studies). Only psychosis and mood disorder risk groups, and only cognitive and affective tasks contributed, in line with only 1/39 available samples including an anxiety risk sample and 2/39 using a reward task (**eFigure 12**).

The sample size was broadly similar across contributing samples (16-30; **eFigure 12**). The percent contribution of each sample was also broadly similar (10.03-25.82% for 5/6 and 0.11% for 1/6 samples; **eFigure 12**). Included and excluded samples differed in mean age (p=0.0061), with included samples having a higher median study-level mean age than excluded samples. No statistically significant differences between included and excluded samples were found in terms of percent female, disorder-risk group distribution, or task domain distribution. Data on sex were not available for three studies.

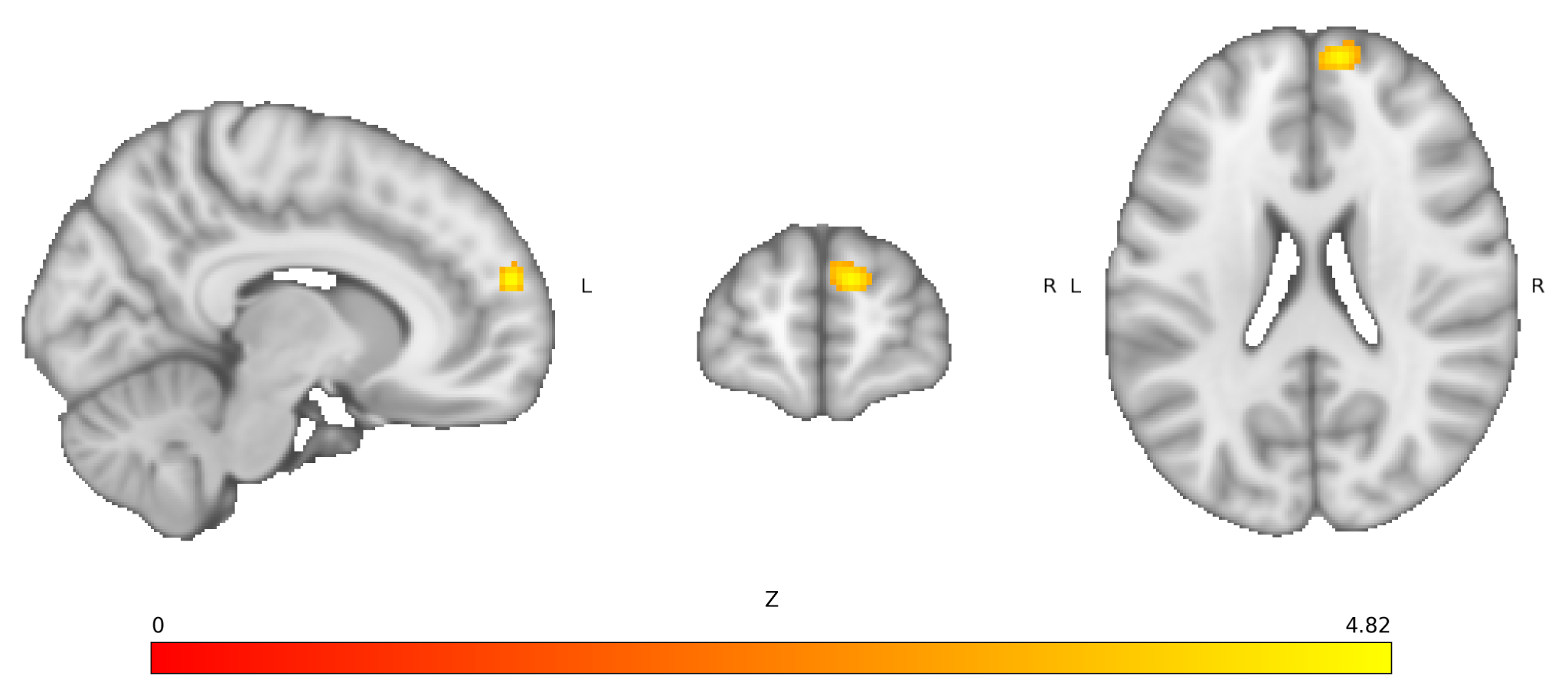

###### eFigure 11: Brain Activation Convergence (p <.05, cFWE) in the FHR Meta-Analysis

The cluster is in the right frontal pole (center: x=10, y=60, z=22).

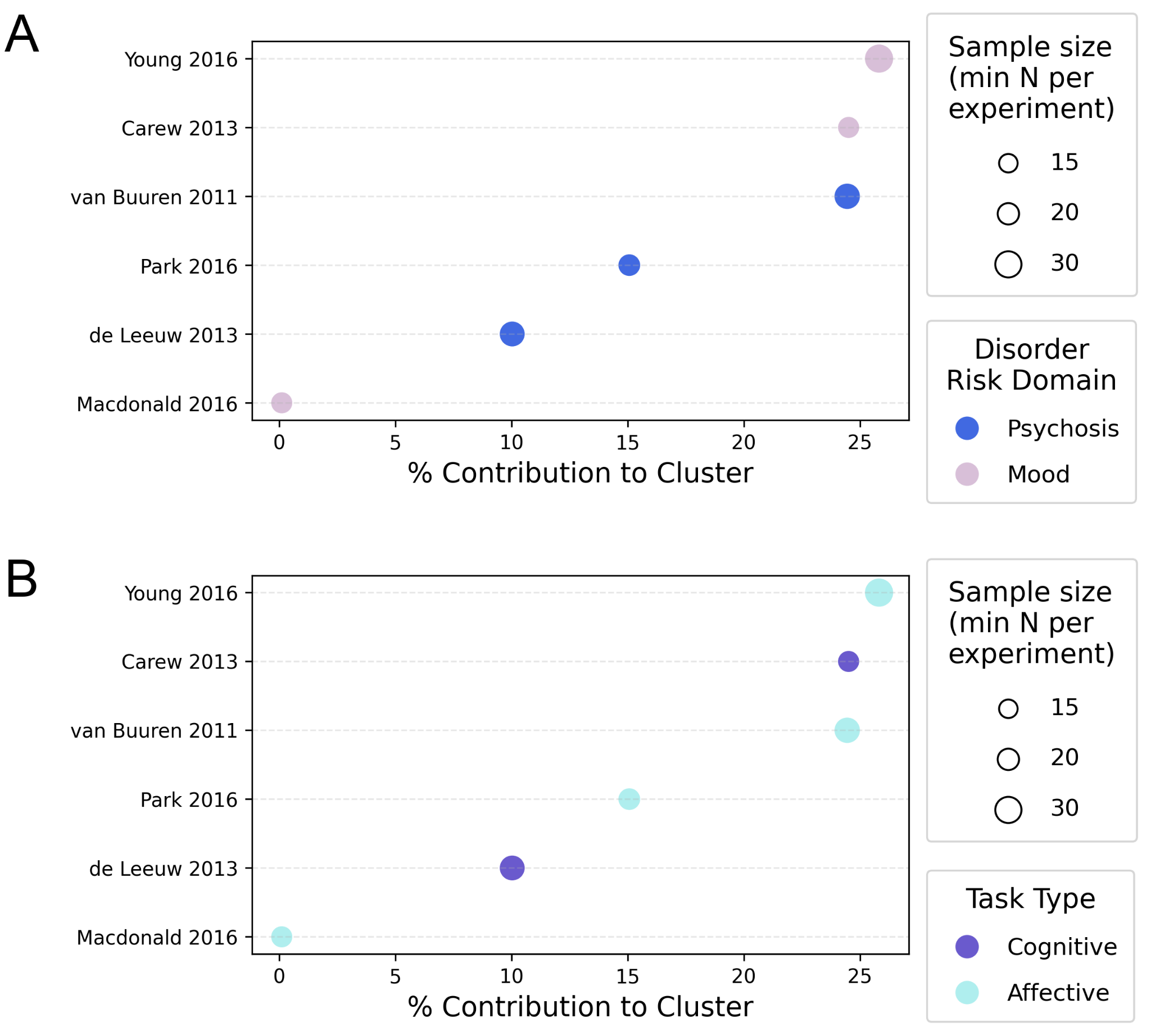
**eFigure 12: (A) Psychosis and Mood Risk Samples and (B) Cognitive and Affective Tasks Contributed to Brain Activation Convergence in the FHR Meta-Analysis**

Sample size here referred to the smaller size across the control and risk sample in each study, which was used for Gaussian kernel weighting in ALE. FHR = familial high-risk.

*PR Sample (cFWE):*

The PR risk sample (64 studies) had two statistically significant clusters (**Figure 3C, eFigure 13**): right anterior insula and OFC (k = 101; 10/64 studies); and left inferior frontal gyrus (k = 203; 20/64 studies). For cluster 1, which included the right anterior insula and OFC, all but one contributing sample (a minor contribution from an anxiety-risk sample) used a psychosis-risk sample and only samples with cognitive and reward tasks contributed. For cluster 2, which included the left inferior frontal gyrus, studies which contributed to these findings spanned all three disorder-risk groups (psychosis, mood, anxiety) and all three task domains (cognitive, reward, affective; **eFigures 14-15**).

For both clusters, the sample size used for Gaussian kernel weighting was consistent across all but one contributing study (cluster 1: 7-40 for all 10 samples, cluster 2: 10-30 for 19/20 and 108 for 1/20 samples; **eFigures 14-15**). The percent contribution of each sample was similar as well (cluster 1: 10.68-18.61% for 6/10 and under 3% for 4/10 samples, cluster 2: 0.08-9.90% for 18/20 and over 12% for 2/20 samples; **eFigures 14-15**). No statistically significant differences between included and excluded samples were found in terms of mean age, percent female, disorder-risk group distribution or task domain distribution. Data on sex were not available for two samples and the sample containing all three task domains was excluded when testing differences in task distribution.

**eFigure 13: Brain Activation Convergence (p <.05, cFWE) in the PR Meta-Analysis**

(A) Cluster 1 is in the right anterior insula and orbitofrontal cortex (center: x=34, y=24, z=-6).

(B) Cluster 2 is in the left inferior frontal gyrus (center: x=-46, y=18, z=14).

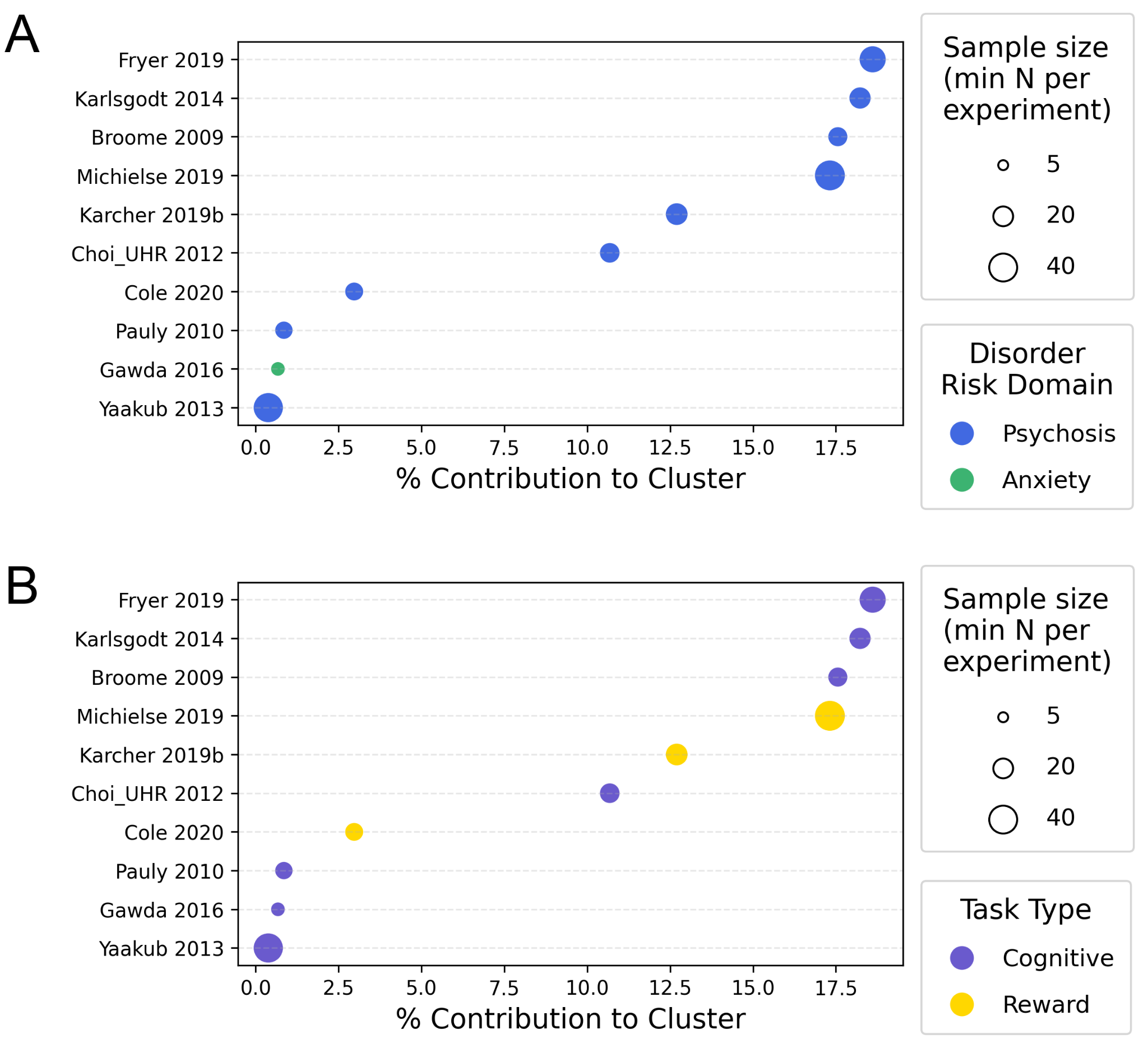
**eFigure 14: (A) Psychosis and Anxiety Risk Samples and (B) Cognitive and Reward Task Domains Contributed to Brain Activation Convergence in Cluster 1 in the PR Meta-Analysis**

Sample size here referred to the smaller size across the control and risk sample in each study, which was used for Gaussian kernel weighting in ALE.

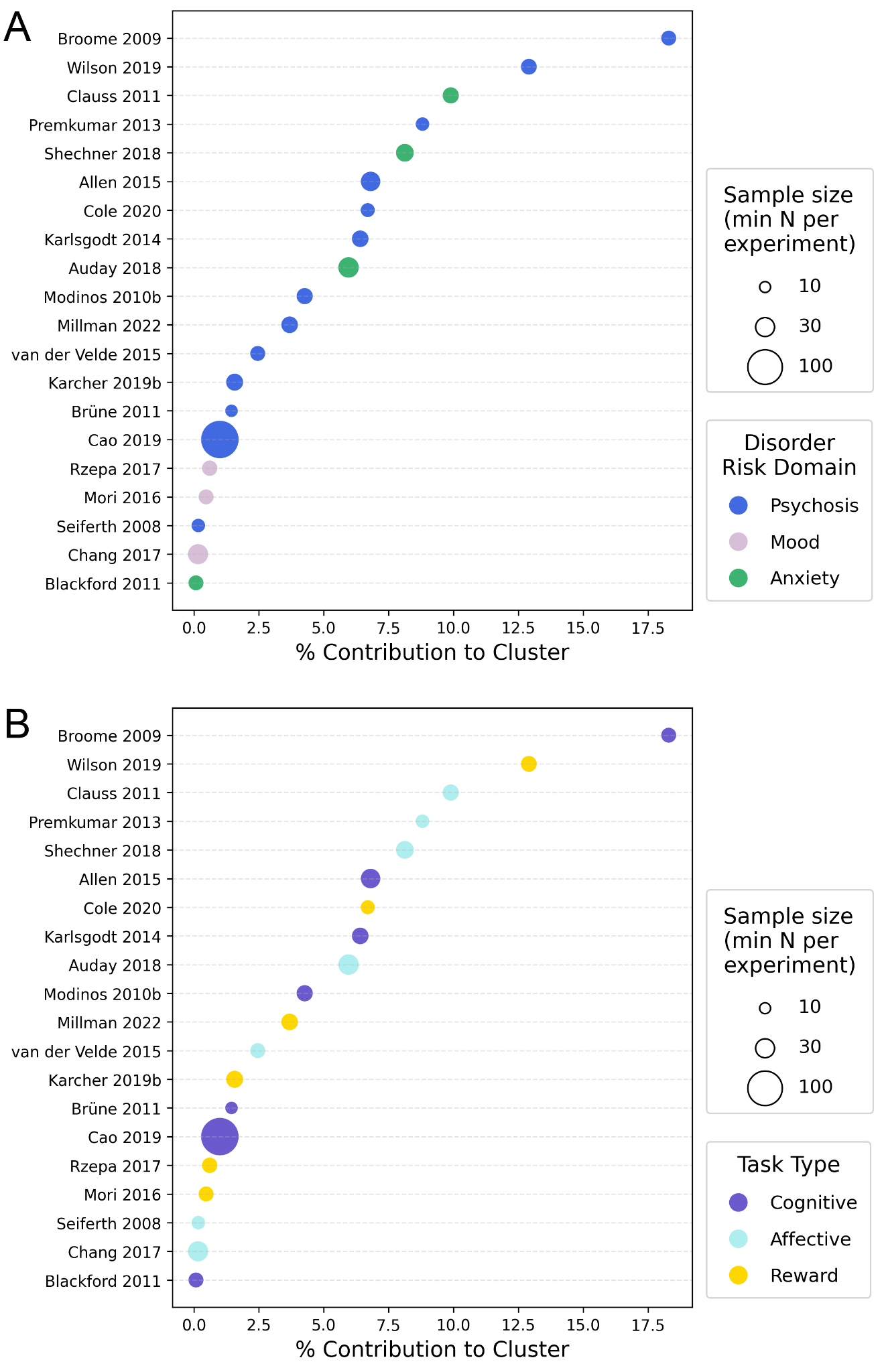

###### eFigure 15: All (A) Disorder-Risk Group Samples and (B) Task Domains Contributed to Brain Activation Convergence in Cluster 2 in the PR Meta-Analysis

Sample size here referred to the smaller size across the control and risk sample in each study, which was used for Gaussian kernel weighting in ALE. PR = phenotypic risk.

*Cluster Overlap Across Individual Meta-Analyses:*

The overall meta-analysis cluster largely overlapped with cluster 3 of the psychosis risk meta-analysis (17 samples shared between the 30/102 samples contributing to the overall meta-analysis and the 19/65 samples contributing to the psychosis risk meta-analysis; **eFigure 16A**). It also substantially overlapped with cluster 2 of the PR meta-analysis (18 samples shared between the 30/102 samples contributing to the overall meta-analysis and the 20/64 samples contributing to the PR meta-analysis; **eFigure 16A**). Cluster 2 of the psychosis risk meta-analysis largely overlapped with cluster 1 of the PR meta-analysis (9 samples shared between the 9/65 samples contributing to the psychosis risk meta-analysis and the 10/64 samples contributing to the PR meta-analysis; **eFigure 16B**).

**eFigure 16. Spatial Overlap of Significant Clusters Across Meta-Analyses.** (A) Overlap in the left lateral prefrontal cortex among the overall meta-analysis across all samples, psychosis-risk meta-analysis Cluster 3, phenotypic-risk meta-analysis Cluster 2, and the overall Risk > Control analysis. (B) Overlap in the right anterior insula between psychosis-risk meta-analysis Cluster 2 and phenotypic-risk meta-analysis Cluster 1. (C) Overlap in the right medial prefrontal cortex among the familial high-risk meta-analysis, the overall Risk > Control analysis, and the overall meta-analysis across all samples. Colored overlays indicate clusters identified in the respective analyses. PR = phenotypic risk; FHR = familial high risk.

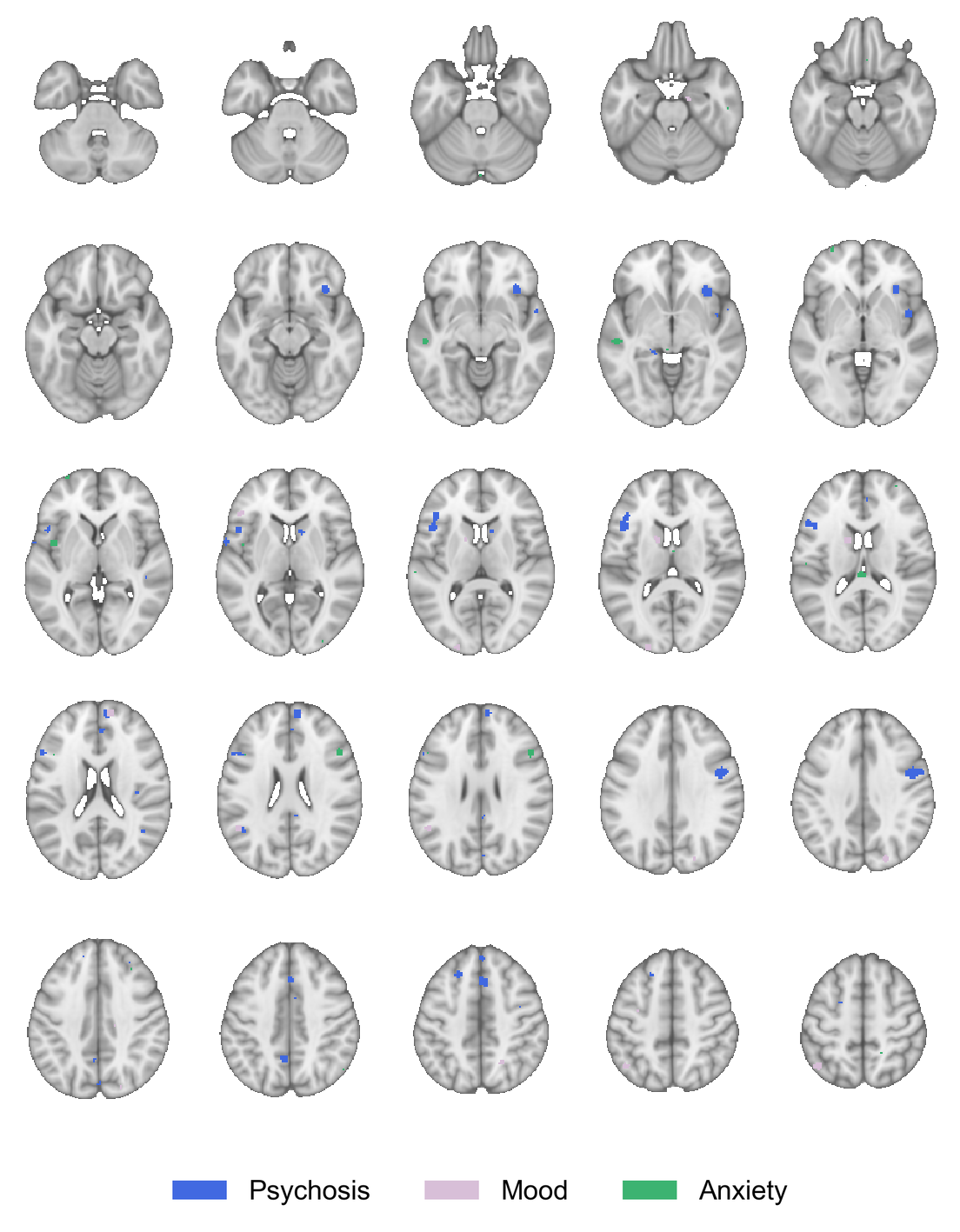
 **eFigure 17. Brain Activation Convergence Across the Three Disorder-Specific Risk Meta-Analyses (p <.001, uncorrected).** Axial brain slices illustrate clusters of convergent activation differences between individuals at elevated psychiatric risk and healthy controls in the psychosis-risk (blue), mood-risk (purple), and anxiety-risk (green) meta-analyses. Psychosis-risk clusters were distributed across default mode network regions (left angular gyrus and precuneus) and frontal regions involved in higher-order processing (left superior frontal gyrus, right paracingulate gyrus, and right frontal pole). Mood-risk clusters involved visual and somatosensory cortices, default mode network regions, the left caudate, and right frontal pole. Anxiety-risk clusters involved frontotemporal and insular regions associated with social-affective processing, as well as the left cerebellum and caudate tail.

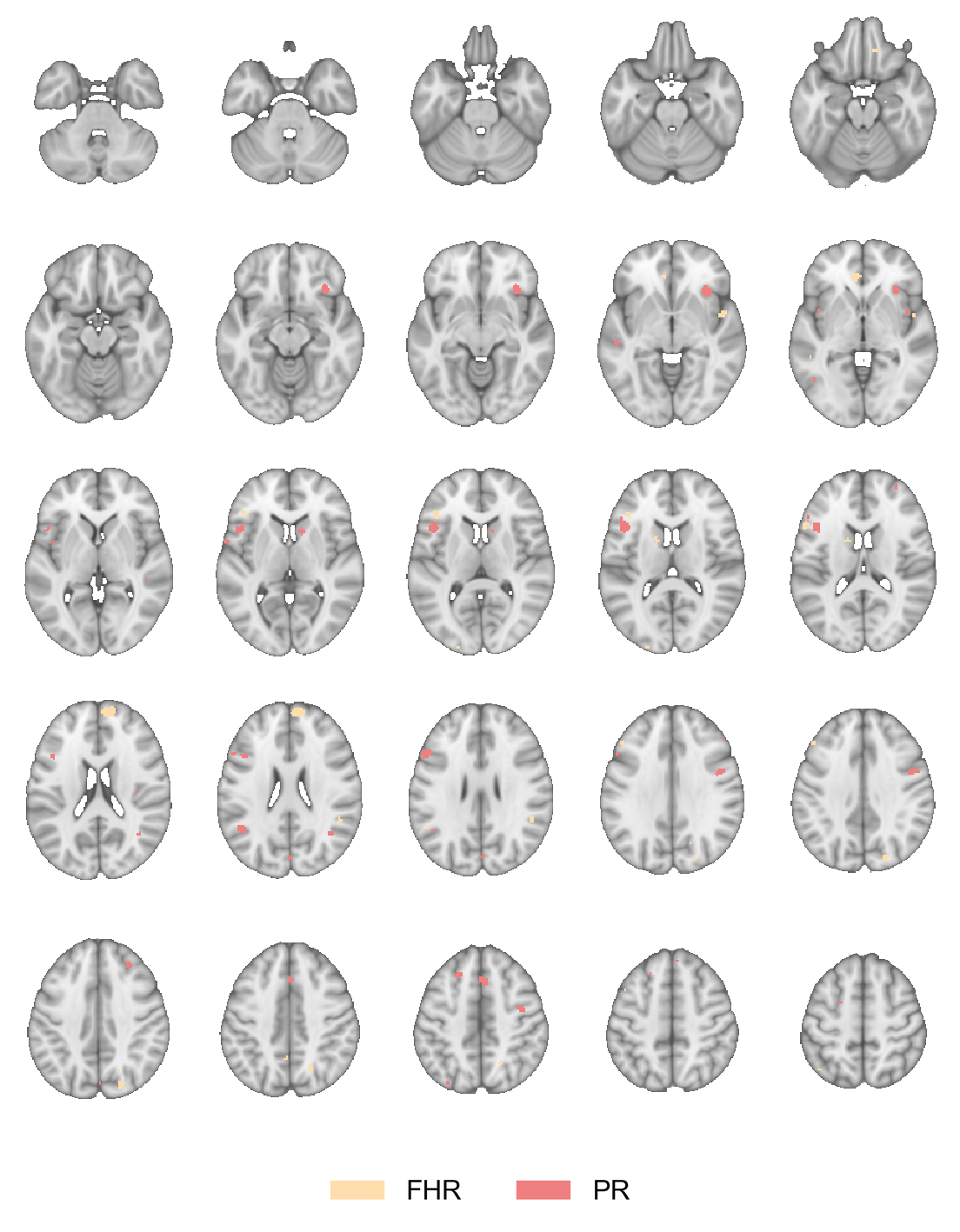

**eFigure 18: Brain Activation Convergence (p <.001, uncorrected) in the Two Ascertainment Group Meta-Analyses.** Axial brain slices illustrate clusters of convergent activation differences between individuals at elevated psychiatric risk and healthy controls in the familial high-risk (FHR; peach) and phenotypic-risk (PR; salmon) meta-analyses. FHR clusters involved default mode network regions (frontal cortex, anterior cingulate, and right angular gyrus), frontoparietal regions (left middle frontal gyrus), bilateral lateral occipital cortex, and right planum polare. PR clusters were more extensively distributed across default mode and frontoparietal network regions, including bilateral angular gyri and frontal cortex, as well as insular, precentral, temporal, and occipital regions.

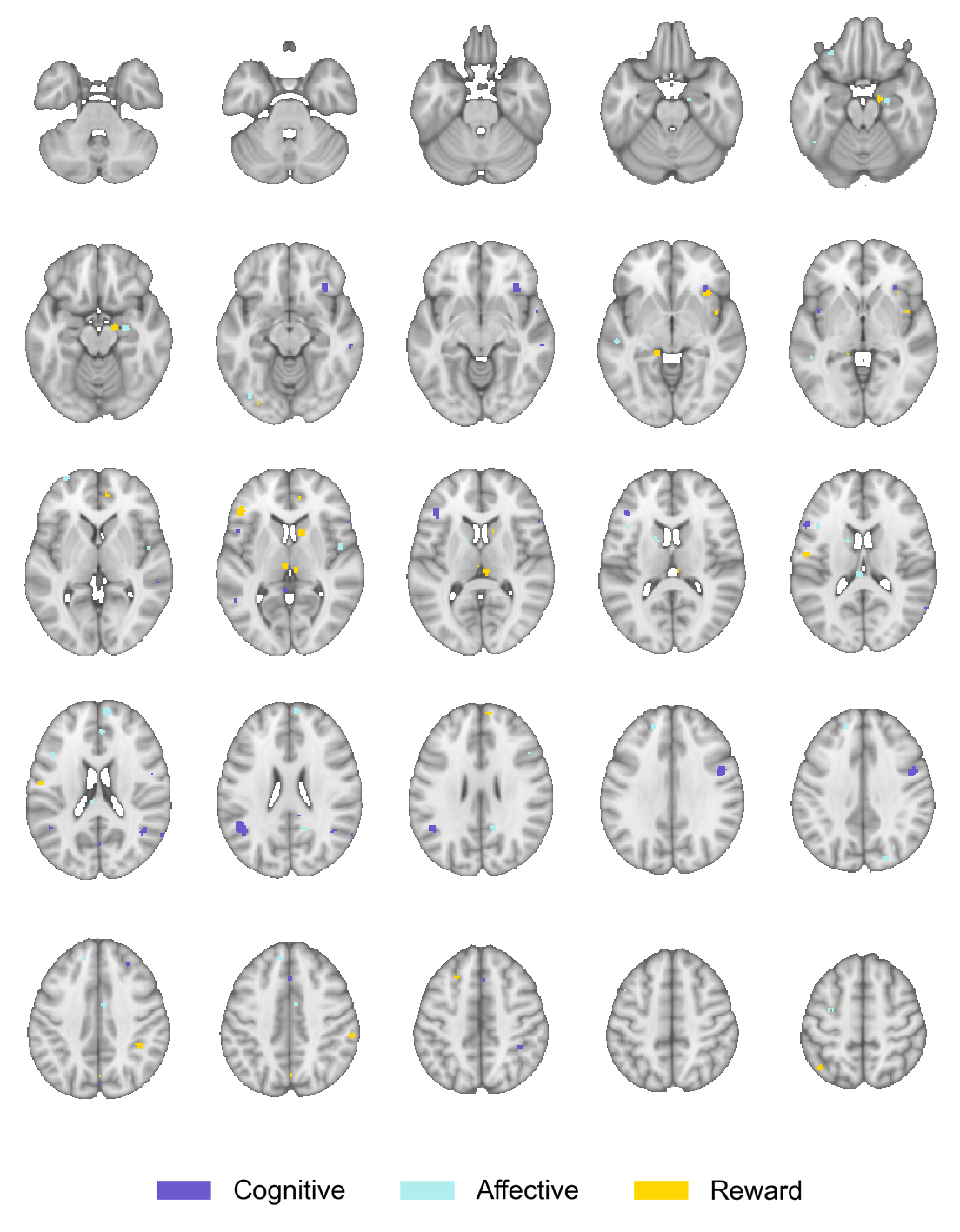

**eFigure 19: Brain Activation Convergence (p <.001, uncorrected) in the Three Task Domain Meta-Analyses.** Axial brain slices illustrate clusters of convergent activation differences between individuals at elevated psychiatric risk and healthy controls in the cognitive (purple), affective (cyan), and reward (yellow) task-domain meta-analyses. Clusters are distributed across frontal, temporal, and parietal regions, with findings across all three task domains. Clusters are displayed at an uncorrected threshold of p <.001.

Sensitivity Analyses

The overall meta-analysis, when repeated with the Gaussian kernel weighted by the combined sample size during ALE instead of the smaller of the risk and control sample sizes, yielded a statistically significant cluster in approximately the same location (center: x=-46, y=19, z=18) but of a smaller size (k=186 voxels). Twenty-five (25) studies contributed to this cluster, which were all shared by the overall meta-analysis cluster. When overlapping samples were merged into one study and the overall meta-analysis was repeated with 96 samples, one statistically significant cluster emerged in the same location (center: x=-46, y=20, z=16) and of approximately the same size (k=328) as the one from the overall meta-analysis. The same samples contributed to the cluster when considering the individual samples that were merged.
